# Cadmium Exposure and Incidence of All-Cause Dementia and Alzheimer’s Disease in US Adults

**DOI:** 10.64898/2026.01.16.26344271

**Authors:** Erika Walker, Yanelli Rodríguez-Carmona, Xin Wang, Bhramar Mukherjee, Laura Arboleda-Merino, Wei Hao, Hiroko Dodge, Roger L. Albin, Henry L. Paulson, Sung Kyun Park, Kelly M. Bakulski

## Abstract

**Introduction:** While longitudinal studies aid in understanding and preventing long-latency disorders like dementia, evidence for cadmium’s role in these conditions is still limited. We evaluated associations between cadmium exposure and incident Alzheimer’s disease (AD) and all-cause dementia in US adults.

**Methods:** National Health and Nutrition Examination Survey (NHANES) III (1988-1994) and continuous NHANES (1999-2016) data were linked with Medicare claims to identify incident AD and dementia cases through 2018. Urinary and/or blood cadmium were measured during NHANES. We used covariate-adjusted, survey-weighted Cox proportional hazard models to evaluate the associations between cadmium exposure biomarkers and AD/dementia over follow-up.

**Results:** In NHANES III (N=6,122), baseline age was 53.9±0.5 years and urinary cadmium was 0.8±0.02 ug/L. Over a follow-up of 20.4±0.3 years, 743 AD and 1,508 all-cause dementia cases occurred. Urinary cadmium was not associated with AD (HR: 1.01, 95% CI: 0.9-1.0) nor all-cause dementia incidence (HR: 1.02, 95% CI: 0.96-1.08). In continuous NHANES (urinary cadmium N=2,833; blood cadmium N=8,038), baseline age was 64.1±0.2 years, urinary cadmium was 0.5±0.03 ug/L, and blood cadmium was 0.6±0.01 ug/L. Over 9.5±0.1 years, 587 AD and 1,260 all-cause dementia cases occurred. Urinary and blood cadmium showed no associations with AD (HR [95% CI]: 1.09 [0.9, 1.4]; 1.06 [0.9, 1.2]) nor all-cause dementia (HR [95% CI]: 1.07 [0.9, 1.3]; 1.06 [0.95, 1.2]).

**Conclusion:** No association between cadmium exposure and dementia incidence was observed. Our null findings should be interpreted while considering potential methodological issues and verified by subsequent studies.

## Introduction

Dementia consists of a group of cognitive function symptoms spanning memory, language, reasoning, and behavioral changes.^1^ In 2019, an estimated 57.2 million people had dementia worldwide, and this number is expected to increase to 83 million by 2030 and to 152.8 million by 2050.^2^ Alzheimer’s disease (AD) is the most common form of dementia, implicated in 60-80% of cases.^1^ Although genetics play a critical role, particularly in early onset AD (<65 years),^3^ the majority of late onset AD (>65 years) is attributed to environmental factors and gene-environment interactions.^4^ Since there are no cures currently available for dementia or AD, prevention is especially important. According to the 2024 Lancet Commission’s dementia prevention report, around 45% of worldwide dementias are attributable to 14 potentially modifiable risk factors. These factors are less education in early life, cardiometabolic risk factors (high low-density lipoprotein cholesterol, physical inactivity, diabetes, smoking, hypertension, obesity, and excessive alcohol consumption), hearing loss, depression and traumatic brain injury in midlife, and social isolation, air pollution and visual loss in later life.^5^ A deeper understanding of modifiable factors is crucial in reducing dementia risk.

Exposure to environmental risk factors, such as cadmium and other non-essential metals, have been identified as hazards in AD development.^6^ Among nonsmoking and nonoccupationally exposed populations, the main exposure source of cadmium is through the diet. After the ingestion of contaminated food, cadmium is absorbed in the intestines and distributed in blood.^7^ Cadmium alters the integrity of the choroid plexus, a component of the blood cerebrospinal fluid barrier,^8^ promoting its accumulation in this structure.^8, 9^ Furthermore, it has been suggested that cadmium can increase the blood-brain barrier permeability.^10^ Cadmium can also reach the nervous tissue, bypassing the blood-brain barrier through the olfactory nervous system.^11, 12^ This is relevant, as cadmium particles can be inhaled through tobacco smoke and ambient air, where they are absorbed by the lungs and enter the bloodstream.^7^ Once in the central nervous system, cadmium induces oxidative stress, neuroinflammation, and apoptosis of neuronal cells, increasing the risk for neurodegenerative diseases.^13, 14^ Although animal studies provide compelling evidence linking cadmium exposures and AD, human studies are essential to elucidate this relationship and establish potential public health prevention strategies.

Current epidemiological evidence points to an association between cadmium exposure and cognitive decline.^15^ Studies on cadmium, dementia, and AD have mostly focused on cognitive function measures^16–25^ and AD mortality.^26–28^ Studies on cadmium and AD mortality have explored this association using the United States (US) National Health and Nutrition Examination Survey (NHANES) and have found that higher blood cadmium was associated with higher AD mortality risk.^26–28^ We are aware of only one study exploring the relation between cadmium and dementia incidence. The Multi-Ethnic Study of Atherosclerosis, which recruited participants from six US study centers, found that one interquartile range increase in log-transformed urinary cadmium concentration at baseline was associated with 17% higher risk for all-cause dementia after a 16.7-year follow-up.^22^ More longitudinal studies are needed to understand the role of environmental exposures, such as cadmium, prior to the onset of dementia. The pathophysiological processes that occur in the development of dementia and AD start long before the appearance of clinical symptoms.^29^ The long latency of dementia suggests that events and exposures that occur before a clinical diagnosis could play a role in the initiation of neurodegeneration. Prospective studies could provide useful information to better prevent and treat these disorders. No longitudinal study on the association between cadmium exposure and AD incidence has been published thus far. Therefore, we evaluated the association between cadmium exposure and the incidence of AD and all-cause dementia in a nationally representative cohort of US adults.

## Methods

### Study sample and data sources

This analysis links data from NHANES with Medicare claims records and the National Death Index (NDI). NHANES is a program of nationally representative cross-sectional surveys of the US civilian population.^30^ NHANES is conducted by the National Center for Health Statistics (NCHS). Data on participant demographics and health status are collected through a series of questionnaires, physical examinations, and laboratory analyses.^30^ We included NHANES III (1988-1994) and continuous NHANES (1999-2016) in this analysis (**Figure 1**).

**Figure 1.**
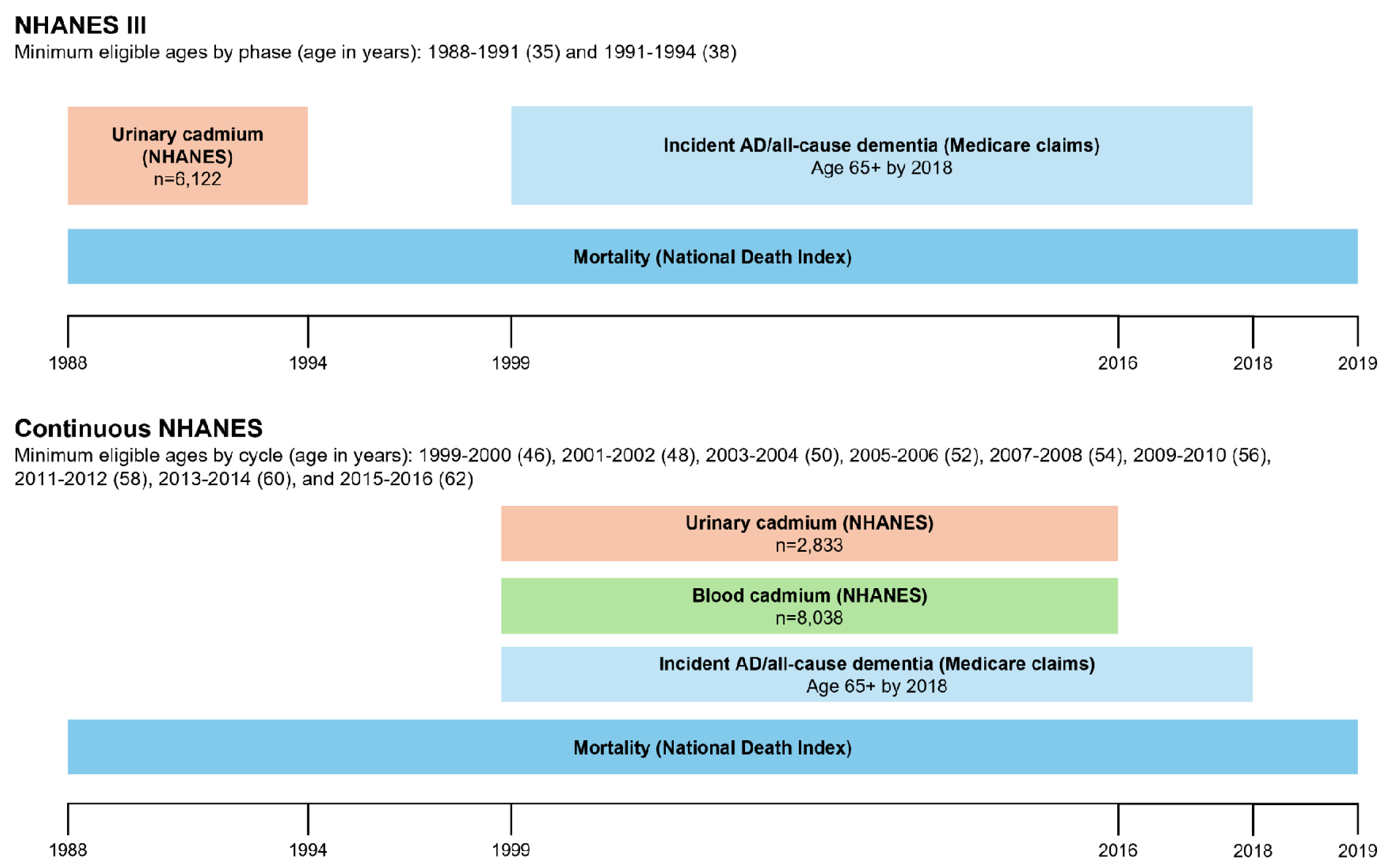
Schematic diagram of the delayed entry of NHANES III and continuous NHANES participants with outcome and follow-up time via Medicare and National Death Index linkage.

Medicare, operated by the Center for Medicare and Medicaid Services (CMS), is the US federal health insurance provider for adults aged 65 and older or people with certain disabilities. NCHS offers linkage between NHANES surveys and Medicare enrollment and claims records.^31^ Participants that provided consent and necessary personal information such as their date of birth and Social Security Number were considered eligible for linkage by NCHS. Full linkage methodology information is available online.^32^ Linked NHANES-Medicare datasets are restricted-use and must be analyzed in federal statistical Research Data Centers (RDCs). To conduct this analysis, we completed a research proposal and a background check review process to obtain Special Sworn Status.

The NDI is a resource operated by the NCHS to provide mortality data for research purposes. While a public-use version NHANES-NDI linkage is available, we instead used the restricted version for more detailed information, including full International Classification of Disease (ICD) cause of death codes and the date of death.^33^ Similar to the Medicare linked files, the restricted dataset is accessible only within an RDC and requires the same application process.

### Cadmium exposure measurement

Cadmium primarily accumulates in the kidneys, with a half-life estimated at 10-15 years.^15, 34^ Measurements from urine therefore represent long-term, cumulative exposure to cadmium, while measurements from blood tend to represent more recent (75 days) exposure.^15^ Urinary cadmium concentrations (ug/L) were measured in both NHANES III and continuous NHANES from spot urine specimens. In NHANES III, urinary cadmium was measured with atomic absorption spectrometry.^35^ The detection limit was 0.01 ug/L. In continuous NHANES, it was measured with inductively coupled plasma mass spectrometry.^36^ The detection limit ranged from 0.036-0.06 ug/L across the cycles. In 1999-2002, measures were corrected for interference from molybdenum oxide.^37, 38^ We removed observations that were corrected to a value of zero. To account for urinary dilution, urinary creatinine (mg/dL) was measured from participant spot urine samples. Continuous NHANES additionally measured whole blood cadmium concentrations (ug/L) using atomic absorption spectrometry from 1999-2002 and inductively coupled plasma mass spectrometry from 2003-2016. Depending on the cycle, the detection limit for blood cadmium ranged from 0.10-0.28 ug/L. In all study years, NHANES imputed measurements below the limit of detection as the limit of detection /√2. We analyzed exposures as continuous variables with log_2_ transformation to interpret model results per doubling of the exposure. As a secondary approach, we also categorized exposures into quartiles based on survey-weighted percentiles and used the lowest quartile as the reference group.

### Outcome assessment

AD and all-cause dementia outcomes were identified primarily from Medicare claims and secondarily from NDI data. We used the Medicare Chronic Conditions segment of the Master Beneficiary Summary Files (1999-2013 and 2014-2018), which contains pre-defined indicator variables for 30 chronic diseases including AD and related dementias.^31, 39^ While CMS uses the term “Alzheimer’s disease and related dementias” in this dataset, we instead used “all-cause dementia” to characterize this outcome. CMS developed these variables using algorithms that searched Medicare fee-for-service claims for the presence of specific ICD codes from ICD-9 or ICD-10 for each condition. A list of the specific ICD-9 and ICD-10 codes used in the outcome definitions is available online through the Medicare Chronic Conditions Data Warehouse,^39^ and a summary of the AD and dementia codes relevant to this analysis is presented in **Table S1**. We created two outcome variables, one for AD incidence and one for all-cause dementia incidence. AD cases are a subset of all-cause dementia cases. We defined the time of event as the date of the first occurrence of an AD- or all-cause dementia-related claim. In instances when participants had claims for both AD and a different non-AD dementia, the all-cause dementia analysis used the outcome that occurred first while the AD analysis used the first AD-specific claim.

To search for additional AD or all-cause dementia cases that did not appear in Medicare claims codes, we also used the NDI Linked Mortality Restricted Use File (1988-2019).^33^ These linked datasets included information on date of death, primary and contributing causes, and the corresponding ICD codes. We searched the causes of death for the same set of ICD codes used in the Medicare definitions to identify additional AD and dementia cases. NDI data were also used to identify dates of death for follow-up of non-cases. Follow-up time was defined as the years between the participant’s NHANES interview and the first identified outcome event for AD/dementia cases, or the date of last follow-up for non-cases (death, last Medicare claim, or transfer to Medicare Advantage).

### Covariate measures

We selected covariates expected to be associated with cadmium exposure and AD/dementia from NHANES datasets containing participant demographics, health behaviors, and examinations. We included baseline age, sex, race and ethnicity, education, poverty-income ratio, smoking status, smoking pack-years, serum cotinine concentration, alcohol consumption, and body mass index (BMI). In NHANES public data, age was top-coded at 80 or 85 depending on the survey year. To reduce potential bias from this truncation, we used participant date of birth as provided in restricted-use files to recode age for this subset. Sex was categorized as male or female. Race and ethnicity were categorized as non-Hispanic White, non-Hispanic Black, Mexican American, other Hispanic (continuous NHANES only), or other non-Hispanic (including multi-racial). Education was categorized as less than high school, completed high school or some college, or completed college and above. Poverty-income ratio was categorized as above or below 1, which represents an income of 100% of the poverty level. Smoking status was categorized from survey responses as never, former, or current smoker. Tobacco pack-years were calculated based on survey responses on average frequency of smoking; for non-smokers, the value was set to zero. Serum cotinine (ng/mL) was measured from participant blood samples. Alcohol consumption was categorized as <1 or ≥1 drink per day, as an average of the past 30 days of reported consumption. BMI was calculated from direct measurements of participant height (m) and weight (kg) during physical examinations.

### Statistical analysis

Participants were included in this analysis if they were at least 65 years old by the end of 2018, were successfully linked to Medicare claims data, did not have AD or dementia claims prior to NHANES participation, and had complete cadmium measurement and covariate data. In NHANES III, n=6,122 participants were included. In continuous NHANES, n=8,038 participants were included with blood cadmium measures and n=2,833 were included with urinary cadmium measures. Due to differences in measurements and sampling weight designs, we analyzed NHANES III and continuous NHANES separately. Analyses were conducted in the secure RDC environment using SAS version 9.4 and R version 4.4.2. Unless noted otherwise, all analyses accounted for the complex survey design, including clustering, stratification and sampling weights, to obtain nationally representative estimates.

We first evaluated descriptive statistics of the study samples by comparing included versus excluded participants with Medicare linkage who were dropped due to missing data. We also compared included participants by outcome statuses (AD versus non-AD and dementia versus non-dementia). Continuous variable means were compared using t-tests and categorical variables were compared using Chi-square tests.

To visualize associations, we plotted unweighted Kaplan-Meier curves of the probability of survival from AD/dementia across low and high cadmium concentrations, divided at the weighted median concentration. To evaluate associations between measures of cadmium exposure and the risks of AD and dementia, we ran separate Cox proportional hazard models for each outcome within each NHANES sample. We presented models based on two forms of the cadmium measurement: log_2_-transformed and quartiles (Q) using Q1 as reference. Models were adjusted for age, sex, race and ethnicity, education, poverty-income ratio, smoking status, pack-years, serum cotinine, alcohol consumption, and BMI. Models for urinary cadmium were further adjusted for urinary creatinine concentration to account for urine dilution. We reported hazard ratios (HR) and 95% confidence intervals (CI).

### Sensitivity analyses

To further explore these associations, we conducted four sensitivity analyses. First, we considered effect modification by age by categorizing participants as less than 65 years and 65 years or older at baseline. We included an effect modification term between cadmium and categorical age, then calculated hazard ratios separately for each age group. To test if residual confounding by age still existed in this model, we also further adjusted for continuous age. Second, we also considered effect modification by sex and generated hazard ratios separately for male and female participants using similar methods as the age model. Third, since cigarette smoking can be a major source of cadmium, we conducted stratified analyses restricted to participants who reported never smoking. Lastly, we conducted Fine-Gray competing risk regression^40^ to account for death due to other causes as a competing outcome. The competing risk models did not account for NHANES survey design due to a lack of software capability. For these models, we reported sub-distribution hazard ratios (SHRs) and 95% CI.

## Results

### Descriptive statistics

Participants in NHANES III (n=6,122) had a mean baseline age of 53.9 years, half were female (53.1%), and 82.5% were Non-Hispanic White. During a mean follow-up period of 20.35 years, 1,508 participants developed all-cause dementia, and 743 participants developed AD (**Table 1**). Participants in continuous NHANES (n=8,038) were on average 64.3 years old at baseline, 54.6% were female (53.1%), and 81.4% were Non-Hispanic White. Over a mean follow-up period of 9.46 years, 1,260 participants developed all-cause dementia, and 587 participants developed AD (**Table 2**). Participants who developed all-cause dementia or AD in both NHANES III and continuous NHANES were more likely to be female, of older age, non-smokers, have lower education levels, lower income, and lower alcohol consumption (**Tables 1, 2**). Only in continuous NHANES, individuals who developed these outcomes were more often Non-Hispanic White, and lower BMI, and lower cotinine levels (**Table 2**).

**Table 1.** Baseline characteristics of included NHANES III participants (N=6,122), overall and stratified by all-cause dementia and by Alzheimer’s disease (AD) outcome status.

|  | All<br>N=6,122 | Non-dementia<br>N=4,614 | All-cause dementia<br>N=1,508 | *p*-value | Non-AD<br>N=5,379 | AD<br>N=743 | *p* -value |
| --- | --- | --- | --- | --- | --- | --- | --- |
| **Continuous variables, mean (SE)** |  |  |  |  |  |  |  |
| Urinary cadmium, ug/L | 0.76 (0.02) | 0.75 (0.02) | 0.83 (0.04) | 0.06 | 0.76 (0.02) | 0.83 (0.05) | 0.21 |
| Follow-up time, years | 20.35 (0.26) | 21.07 (0.27) | 17.15 (0.28) | **<0.0001** | 20.85 (0.27) | 17.49 (0.38) | **<0.0001** |
| Age, years | 53.86 (0.46) | 51.45 (0.45) | 64.61 (0.47) | **<0.0001** | 52.76 (0.47) | 64.85 (0.51) | **<0.0001** |
| Pack-years | 14.29 (0.47) | 14.15 (0.53) | 14.94 (0.92) | 0.46 | 14.28 (0.48) | 14.38 (1.21) | 0.94 |
| Serum cotinine, ng/mL | 70.47 (2.65) | 74.37 (2.69) | 53.49 (5.88) | **0.0009** | 72.18 (2.73) | 53.92 (8.58) | **0.04** |
| Body mass index, kg/m<sup>2</sup> | 27.36 (0.14) | 27.36 (0.16) | 27.36 (0.24) | 0.99 | 27.36 (0.15) | 27.37 (0.32) | 0.96 |
| Urinary creatinine, mg/dL | 113.78 (1.64) | 116.43 (1.88) | 102.21 (2.28) | **<0.0001** | 115.07 (1.74) | 101.24 (3.69) | **0.002** |
| **Categorical variables, n (%)** |  |  |  |  |  |  |  |
| Sex |  |  |  | **<0.0001** |  |  | **<0.0001** |
| Male | 2,797 (46.88%) | 2,170 (48.71%) | 627 (38.70%) |  | 2,506 (48.01%) | 291 (35.62%) |  |
| Female | 3,325 (53.12%) | 4,444 (51.29%) | 881 (61.30%) |  | 2,873 (51.99%) | 452 (64.38%) |  |
| Race/ethnicity |  |  |  | 0.16 |  |  | 0.29 |
| Non-Hispanic White | 3,130 (82.54%) | 2,272 (82.31%) | 858 (83.57%) |  | 2,703 (82.43%) | 427 (83.63%) |  |
| Non-Hispanic Black | 1,398 (8.16%) | 1,075 (7.99%) | 323 (8.91%) |  | 1,239 (8.09%) | 159 (8.77%) |  |
| Mexican American | 1,352 (3.37%) | 1,069 (3.53%) | 283 (2.65%) |  | 1,220 (3.49%) | 132 (2.14%) |  |
| Other | 242 (5.93%) | 198 (6.17%) | 44 (4.87%) |  | 217 (5.98%) | 25 (5.45%) |  |
| Education |  |  |  | **<0.0001** |  |  | **<0.0001** |
| Less than high school | 2,491 (24.54%) | 1,703 (21.54%) | 788 (37.87%) |  | 2,107 (23.20%) | 384 (37.75%) |  |
| High school graduate | 2,743 (52.36%) | 2,183 (53.64%) | 560 (46.66%) |  | 2,460 (52.82%) | 283 (47.78%) |  |
| College and above | 888 (23.11%) | 728 (24.82%) | 160 (15.47%) |  | 812 (23.98%) | 76 (14.47%) |  |
| Income-poverty ratio |  |  |  | **<0.0001** |  |  | **0.0009** |
| ≤ 1 | 1,077 (8.34%) | 753 (7.41%) | 324 (12.46%) |  | 921 (7.97%) | 156 (11.99%) |  |
| >1 | 5,045 (91.66%) | 3,861 (92.59%) | 1,184 (87.54%) |  | 4,458 (92.03%) | 587 (88.01%) |  |
| Smoking status |  |  |  | **0.0004** |  |  | 0.09 |
| Never | 2,896 (43.86%) | 2,123 (42.92%) | 773 (48.04%) |  | 2,508 (43.42%) | 388 (48.17%) |  |
| Former | 1,923 (33.45%) | 1,433 (32.98%) | 490 (35.54%) |  | 1,676 (33.34%) | 247 (34.56%) |  |
| Current | 1,303 (22.69%) | 1,058 (24.10%) | 245 (16.42%) |  | 1,195 (23.24%) | 108 (17.27%) |  |
| Alcohol consumption |  |  |  | **0.01** |  |  | 0.049 |
| <1 drink/day | 5,413 (87.03%) | 4,038 (86.43%) | 1,375 (89.62%) |  | 4,733 (86.68%) | 680 (90.41%) |  |
| ≥1 drink/day | 709 (12.97%) | 576 (13.57%) | 133 (10.38%) |  | 646 (13.32%) | 63 (9.59%) |  |

Note: Survey weighted means and standard errors were calculated for continuous variables. Unweighted frequencies and survey weighted percentages were calculated for categorical variables. Survey weighted t-tests were used to test continuous variables, and survey weighted Chi-square test was used for categorical variables.

**Table 2.** Baseline characteristics of included continuous NHANES participants (N=8,038), overall and stratified by all-cause dementia and related dementias and Alzheimer’s disease (AD) outcome status, cadmium analysis.

|  | All<br>N=8,038 | Non-dementia<br>N=6,778 | All-cause dementia<br>N=1,260 | *p*-value | Non-AD<br>N=7,451 | AD<br>N=587 | *p*-value |
| --- | --- | --- | --- | --- | --- | --- | --- |
| **Continuous variables, mean (SE)** |  |  |  |  |  |  |  |
| Blood cadmium, ug/L | 0.55 (0.01) | 0.54 (0.01) | 0.57 (0.01) | 0.06 | 0.55 (0.01) | 0.57 (0.03) | 0.39 |
| Urinary cadmium,ug/L | 0.49 (0.03) | 0.48 (0.03) | 0.54 (0.03) | 0.14 | 0.49 (0.03) | 0.48 (0.04) | 0.94 |
| Follow-up time, years | 9.46 (0.12) | 9.66 (0.13) | 7.98 (0.20) | **<0.0001** | 9.74 (0.13) | 8.53 (0.22) | **<0.0001** |
| Age, years | 64.13 (0.19) | 63.01 (0.20) | 72.18 (0.33) | **<0.0001** | 63.60 (0.19) | 73.21 (0.48) | **<0.0001** |
| Pack-years | 14.03 (0.37) | 14.02 (0.39) | 14.07 (0.98) | 0.96 | 14.12 (0.38) | 12.42 (1.32) | 0.21 |
| Serum cotinine, ng/mL | 45.38 (1.78) | 46.55 (1.93) | 37.34 (3.75) | **0.03** | 46.39 (1.86) | 29.13 (4.35) | **0.0003** |
| Body mass index, kg/m<sup>2</sup> | 28.78 (0.09) | 28.93 (0.10) | 27.74 (0.17) | **<0.0001** | 28.89 (0.10) | 27.07 (0.25) | **<0.0001** |
| Urinary creatinine, mg/dL | 106.24 (0.98) | 106.45 (0.97) | 104.81 (2.63) | 0.53 | 106.71 (0.99) | 98.65 (3.45) | **0.02** |
| **Categorical variables, n (%)** |  |  |  |  |  |  |  |
| Sex |  |  |  | **<0.0001** |  |  | **<0.0001** |
| Male | 3,822 (45.39%) | 3,267 (46.24%) | 555 (39.34%) |  | 3,588 (46.06%) | 234 (33.99%) |  |
| Female | 4,216 (54.61%) | 3,511 (53.76%) | 705 (60.66%) |  | 3,863 (53.94%) | 353 (66.01%) |  |
| Race/ethnicity |  |  |  | **0.01** |  |  | **0.02** |
| Non-Hispanic White | 4,618 (81.71%) | 3,787 (81.37%) | 831 (84.14%) |  | 4,222 (81.48%) | 396 (85.65%) |  |
| Non-Hispanic Black | 1,403 (7.51%) | 1,199 (7.41%) | 204 (8.19%) |  | 1,323 (7.54%) | 80 (7.02%) |  |
| Mexican American | 1,168 (3.40%) | 1,020 (3.50%) | 148 (2.67%) |  | 1,092 (3.43%) | 76 (2.87%) |  |
| Other Hispanic | 488 (3.35%) | 444 (3.51%) | 44 (2.14%) |  | 469 (3.45%) | 19 (1.62%) |  |
| Other | 361 (4.04%) | 328 (4.20%) | 33 (2.85%) |  | 345 (4.11%) | 16 (2.83%) |  |
| Education |  |  |  | **<0.0001** |  |  | **<0.0001** |
| Less than high school | 2,300 (17.10%) | 1,834 (15.71%) | 466 (27.07%) |  | 2,076 (16.45%) | 224 (28.06%) |  |
| High school graduate | 3,945 (53.43%) | 3,353 (53.44%) | 592 (53.34%) |  | 3,675 (53.44%) | 270 (53.25%) |  |
| College and above | 1,793 (29.47%) | 1,591 (30.85%) | 202 (19.60%) |  | 1,700 (30.11%) | 93 (18.69%) |  |
| Income-poverty ratio |  |  |  | **<0.0001** |  |  | **0.0004** |
| ≤ 1 | 1,191 (8.32%) | 965 (7.68%) | 226 (12.93%) |  | 1,092 (8.07%) | 99 (12.58%) |  |
| >1 | 6,847 (91.68%) | 5,813 (92.32%) | 1,034 (87.07%) |  | 6,359 (91.93%) | 488 (87.42%) |  |
| Smoking status |  |  |  | **0.02** |  |  | **0.01** |
| Never | 4,087 (50.13%) | 3,406 (49.61%) | 681 (53.85%) |  | 3,756 (49.77%) | 331 (56.21%) |  |
| Former | 2,739 (34.75%) | 2,308 (34.80) | 431 (34.40%) |  | 2,531 (34.77%) | 208 (34.52%) |  |
| Current | 1,212 (15.12%) | 1,064 (15.59%) | 148 (11.74%) |  | 1,164 (15.46%) | 48 (9.28%) |  |
| Alcohol consumption |  |  |  | **0.005** |  |  | **0.002** |
| <1 drink/day | 7,023 (85.78%) | 5,886 (85.23%) | 1,137 (89.58%) |  | 6,483 (85.41%) | 540 (91.80%) |  |
| ≥1 drink/day | 1,015 (14.22%) | 892 (14.77%) | 123 (10.42%) |  | 968 (14.59%) | 47 (8.20%) |  |

Note: Survey weighted means and standard errors were calculated for continuous variables. Unweighted frequencies and survey weighted percentages were calculated for categorical variables. Survey weighted t-tests were used to test continuous variables, and survey weighted Chi-square test was used for categorical variables.
\* Urinary cadmium measurements were available in a subset of 2,833 participants (407 all-cause dementia and 2,426 non-dementia; 2,650 AD and 183 non-AD.)

Participants excluded from the NHANES III analytical sample tended to be female, older, non-White, current or former smokers, have lower income and lower education levels. No differences were observed for urinary cadmium concentrations (**Table S2**). On the other hand, the participants from continuous NHANES excluded from the analysis had higher blood cadmium concentrations, were older, non-White, had less education level, lower income, and lower BMI, and were more likely to be former or current smokers and had more pack years. Urinary cadmium concentrations were similar between our sample and excluded participants (**Table S2**).

### NHANES III: Cadmium and AD/all-cause dementia

The unadjusted Kaplan-Meier curves in NHANES III showed that participants with urinary cadmium concentrations above the median had similar AD and all-cause dementia survival rate during the follow-up, compared to those with urinary cadmium concentrations below the median (**Figure S1**). Using multivariable adjusted survey-weighted Cox regression proportional hazards models, no associations were observed between urinary cadmium and incident AD nor all-cause dementia (**Table 3**). As examples of these null observations, a doubling of urinary cadmium was associated with 1% hazard for AD (HR: 1.01; 95% CI: 0.92, 1.10) and 2% hazard for all-cause dementia (HR: 1.02; 95% CI: 0.96, 1.08).

**Table 3.** Associations between urinary cadmium and incident Alzheimer’s disease (AD) and all-cause dementia in NHANES III (N=6,122)

|  | Quartiles of urinary cadmium |  |  |  |  | Continuous urinary cadmium |  |
| --- | --- | --- | --- | --- | --- | --- | --- |
|  | Q1 | Q2<br>HR (95% CI) | Q3<br>HR (95% CI) | Q4<br>HR (95% CI) | *p*-trend | Per doubling<br>HR (95% CI) | *p* |
| **AD** |  |  |  |  |  |  |  |
| Range (ug/L) | 0.01-0.23 | 0.24-0.50 | 0.51-0.97 | 0.98-12.26 |  |  |  |
| Cases / Quartile | 154/1,305 | 181/1,487 | 204/1,644 | 204/1,686 |  |  |  |
| Estimates | Ref | 0.94 (0.68, 1.30) | 0.77 (0.54, 1.10) | 0.95 (0.60, 1.49) | 0.56 | 1.01 (0.92, 1.10) | 0.83 |
| **All-cause dementia** |  |  |  |  |  |  |  |
| Range (ug/L) | 0.01-0.23 | 0.24-0.50 | 0.51-0.97 | 0.98-12.26 |  |  |  |
| Cases / Quartile | 320/1,305 | 365/1,487 | 406/1,644 | 417/1,686 |  |  |  |
| Estimates | Ref | 0.99 (0.82, 1.20) | 0.90 (0.72, 1.11) | 1.01 (0.74, 1.36) | 0.80 | 1.02 (0.96, 1.08) | 0.60 |

Notes: Quartiles (Q) of cadmium concentrations are based on weighted distributions.
All models were adjusted for age, sex, race/ethnicity, education, income-poverty ratio, smoking status, pack-year, serum cotinine, alcohol consumption, body mass index, and urinary creatinine.

### Continuous NHANES: Cadmium and AD/all-cause dementia

In continuous NHANES, the unadjusted Kaplan-Meier curves showed that participants with blood cadmium concentrations above the medians had lower AD and all-cause dementia survival rate during the follow-up, compared to those with blood cadmium concentrations below the median (**Figure S2**). AD and all-cause dementia survival rates were comparable for participants with urinary cadmium concentrations above and below the median (**Figure S3**). Similar to what we observed in NHANES III, in continuous NHANES no associations were observed between urinary cadmium, blood cadmium and incidence of AD or all-cause dementia (**Table 4**).

**Table 4.**
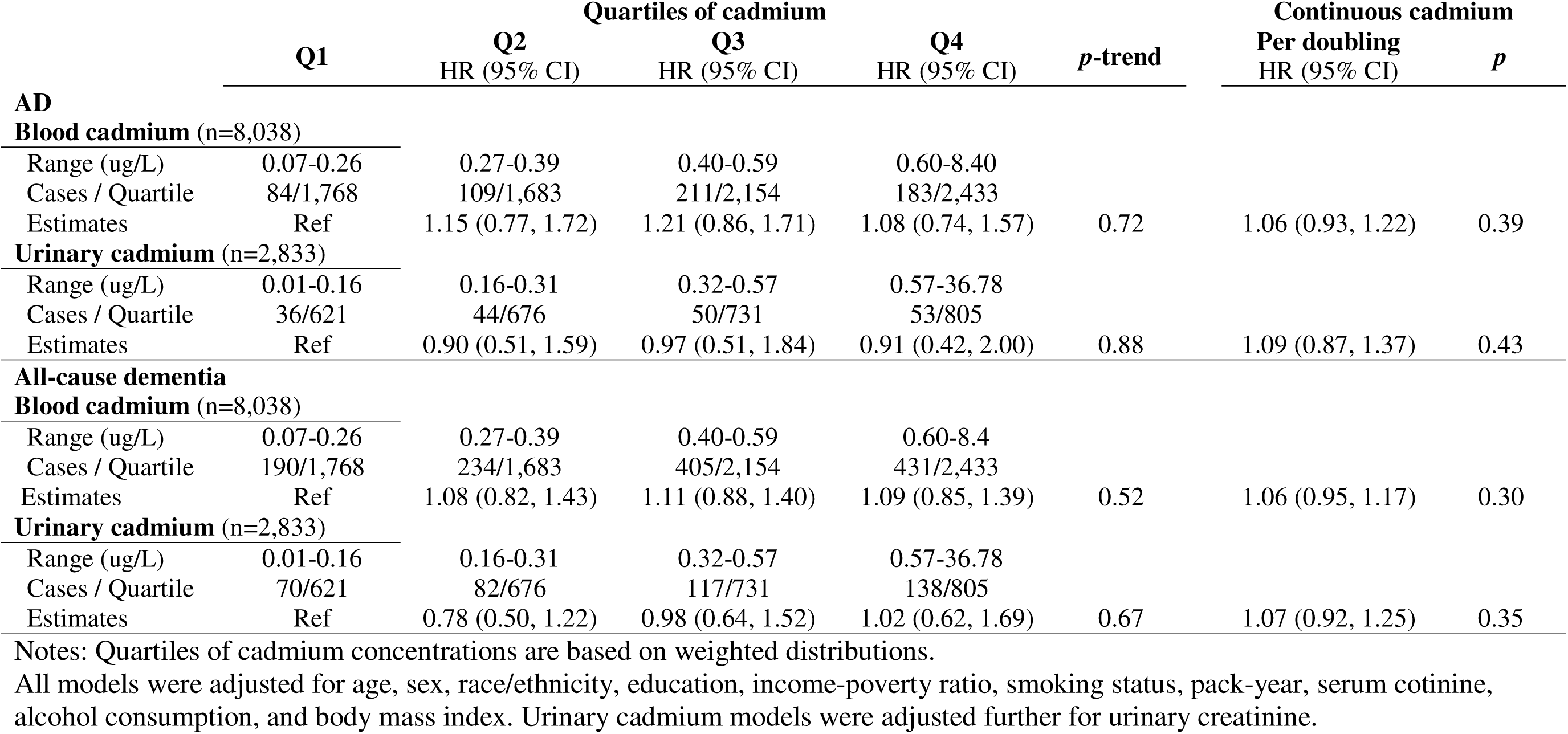
Associations between cadmium and incident Alzheimer’s disease (AD) and all-cause dementia in continuous NHANES.

### Sensitivity analyses

In NHANES III, we observed positive associations between urinary cadmium, AD, and all-cause dementia only in participants younger than 65 years of age at the time of exposure measurement (**Table 5**). The highest urinary cadmium quartile had 78% (HR: 1.78; 95% CI: 1.04, 3.05) higher hazard of incident AD, relative to the lowest cadmium quartile. Similarly, the highest urinary cadmium quartile had 93% (HR: 1.93; 95% CI: 1.35, 2.75) higher hazard for incident all-cause dementia, when compared to the lowest cadmium quartile. Similar findings were observed for continuous urinary cadmium, where each doubling of urinary cadmium was associated with 16% (HR: 1.16; 95% CI: 1.04, 1.30) higher hazard for incident AD and 17% (HR: 1.17; 95% CI: 1.09, 1.26) higher hazard for incident all-cause dementia. However, these age-stratified associations were no longer statistically significant after further adjusting for continuous age (**Table 5**).

**Table 5.** Effect modification of associations of urinary cadmium exposure with incident Alzheimer’s disease (AD) and all-cause dementia by age group in NHANES III (N=6,122)

|  | Quartiles of urinary cadmium |  |  |  |  | Continuous urinary cadmium |  |
| --- | --- | --- | --- | --- | --- | --- | --- |
|  | Q1 | Q2<br>HR (95% CI) | Q3<br>HR (95% CI) | Q4<br>HR (95% CI) | *p* for interaction | Per doubling<br>HR (95% CI) | *p* for interaction |
| **AD** |  |  |  |  |  |  |  |
| *Unadjusted for continuous age* |  |  |  |  |  |  |  |
| Age <65 (n=4,193) | Ref | 1.33 (0.85, 2.09) | 1.01 (0.54, 1.87) | 1.78 (1.04, 3.05) | 0.35 | 1.16 (1.04, 1.30) | 0.62 |
| Age 65+ (n=1,929) | Ref | 1.02 (0.62, 1.69) | 1.22 (0.76, 1.97) | 1.44 (0.78, 2.69) |  | 1.12 (0.96, 1.30) |  |
| *Adjusted for continuous age* |  |  |  |  |  |  |  |
| Age <65 (n=4,193) | Ref | 0.93 (0.60, 1.44) | 0.55 (0.31, 0.97) | 0.79 (0.47, 1.36) | 0.14 | 0.99 (0.90, 1.09) | 0.59 |
| Age 65+ (n=1,929) | Ref | 0.95 (0.58, 1.54) | 1.00 (0.68, 1.46) | 1.03 (0.59, 1.78) |  | 1.03 (0.90, 1.17) |  |
| **All-cause dementia** |  |  |  |  |  |  |  |
| *Unadjusted for continuous age* |  |  |  |  |  |  |  |
| Age <65 (n=4,193) | Ref | 1.28 (0.89, 1.83) | 1.39 (0.94, 2.05) | 1.93 (1.35, 2.75) | 0.74 | 1.17 (1.09, 1.26) | 0.33 |
| Age 65+ (n=1,929) | Ref | 1.15 (0.88, 1.50) | 1.17 (0.84, 1.64) | 1.47 (0.97, 2.24) |  | 1.11 (0.99, 1.23) |  |
| *Adjusted for continuous age* |  |  |  |  |  |  |  |
| Age <65 (n=4,193) | Ref | 0.93 (0.67, 1.29) | 0.81 (0.55, 1.19) | 0.95 (0.64, 1.41) | 0.89 | 1.01 (0.94, 1.08) | 0.95 |
| Age 65+ (n=1,929) | Ref | 1.04 (0.81, 1.34) | 0.96 (0.74, 1.25) | 1.01 (0.69, 1.50) |  | 1.01 (0.92, 1.11) |  |

All models were adjusted for categorical age, sex, race/ethnicity, education, income-poverty ratio, smoking status, pack-year, serum cotinine, alcohol consumption, body mass index, and urinary creatinine. Multiplicative interaction terms between cadmium and categorical age were also included in the models.

On the other hand, for participants 65 and older in continuous NHANES, blood cadmium was associated with both higher risk for AD and all-cause dementia incidence (**Table 6**). We observed a log-linear association between the blood cadmium quartiles and AD (in Q3 HR=1.47; 95% CI: 1.03, 2.10, and in Q4 HR=1.48; 95% CI: 1.05, 2.08), and all-cause dementia incidence (in Q3 HR =1.43; 95% CI: 1.11, 1.84; and in Q4 HR= 1.50; 95% CI: 1.13, 1.98). Similarly, each doubling of continuous blood cadmium in people 65 and over was associated with 21% (HR: 1.21; 95% CI: 1.05, 1.38) higher hazard for incident AD and 18% (HR: 1.18; 95% CI: 1.02, 1.46) higher hazard for incident all-cause dementia. The only association we observed in continuous NHANES for people aged <65 was between continuous blood cadmium and all-cause dementia (HR: 1.22; 95%: 1.02, 1.46). No statistically significant associations were observed after adjusting for continuous age (**Table 6**).

**Table 6.**
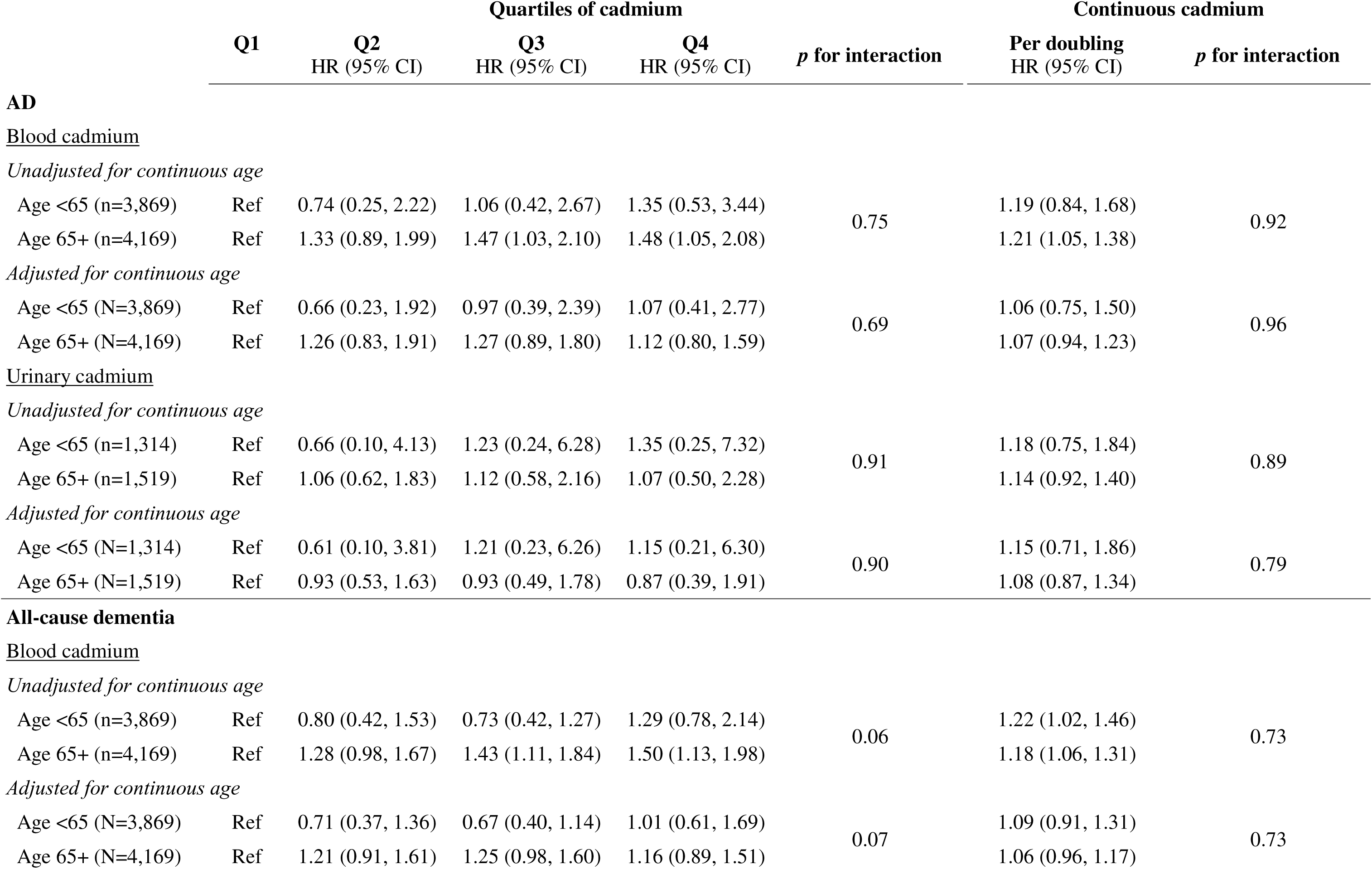
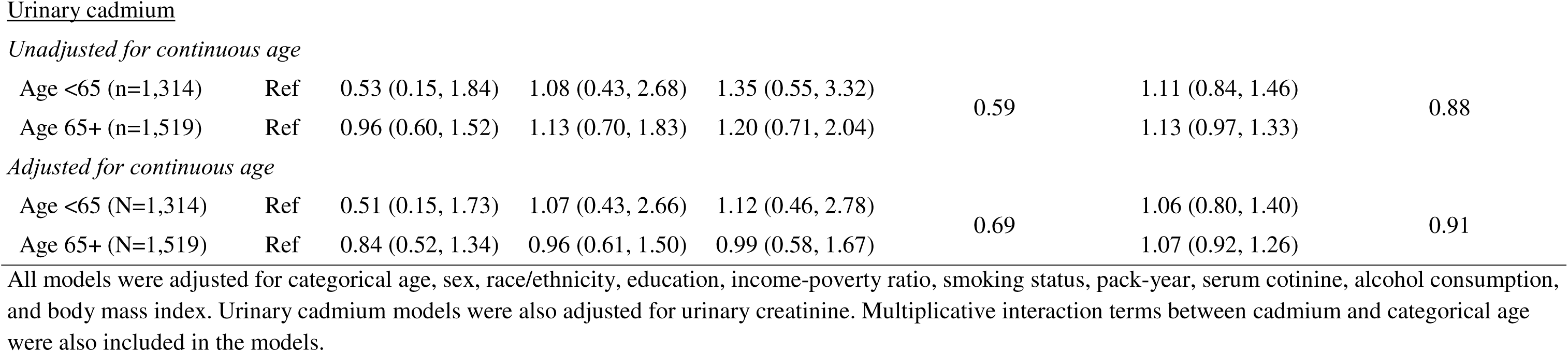
Effect modification of associations between cadmium exposure with incident Alzheimer’s disease (AD) and all cause dementia by age in continuous NHANES.

|  | Quartiles of cadmium |  |  |  |  | Continuous cadmium |  |
| --- | --- | --- | --- | --- | --- | --- | --- |
|  | Q1 | Q2<br>HR (95% CI) | Q3<br>HR (95% CI) | Q4<br>HR (95% CI) | *p* for interaction | Per doubling<br>HR (95% CI) | *p* for interaction |
| **AD** |  |  |  |  |  |  |  |
| <u>Blood cadmium</u> |  |  |  |  |  |  |  |
| *Unadjusted for continuous age* |  |  |  |  |  |  |  |
| Age <65 (n=3,869) | Ref | 0.74 (0.25, 2.22) | 1.06 (0.42, 2.67) | 1.35 (0.53, 3.44) | 0.75 | 1.19 (0.84, 1.68) | 0.92 |
| Age 65+ (n=4,169) | Ref | 1.33 (0.89, 1.99) | 1.47 (1.03, 2.10) | 1.48 (1.05, 2.08) |  | 1.21 (1.05, 1.38) |  |
| *Adjusted for continuous age* |  |  |  |  |  |  |  |
| Age <65 (N=3,869) | Ref | 0.66 (0.23, 1.92) | 0.97 (0.39, 2.39) | 1.07 (0.41, 2.77) | 0.69 | 1.06 (0.75, 1.50) | 0.96 |
| Age 65+ (N=4,169) | Ref | 1.26 (0.83, 1.91) | 1.27 (0.89, 1.80) | 1.12 (0.80, 1.59) |  | 1.07 (0.94, 1.23) |  |
| <u>Urinary cadmium</u> |  |  |  |  |  |  |  |
| *Unadjusted for continuous age* |  |  |  |  |  |  |  |
| Age <65 (n=1,314) | Ref | 0.66 (0.10, 4.13) | 1.23 (0.24, 6.28) | 1.35 (0.25, 7.32) | 0.91 | 1.18 (0.75, 1.84) | 0.89 |
| Age 65+ (n=1,519) | Ref | 1.06 (0.62, 1.83) | 1.12 (0.58, 2.16) | 1.07 (0.50, 2.28) |  | 1.14 (0.92, 1.40) |  |
| *Adjusted for continuous age* |  |  |  |  |  |  |  |
| Age <65 (N=1,314) | Ref | 0.61 (0.10, 3.81) | 1.21 (0.23, 6.26) | 1.15 (0.21, 6.30) | 0.90 | 1.15 (0.71, 1.86) | 0.79 |
| Age 65+ (N=1,519) | Ref | 0.93 (0.53, 1.63) | 0.93 (0.49, 1.78) | 0.87 (0.39, 1.91) |  | 1.08 (0.87, 1.34) |  |
| **All-cause dementia** |  |  |  |  |  |  |  |
| <u>Blood cadmium</u> |  |  |  |  |  |  |  |
| *Unadjusted for continuous age* |  |  |  |  |  |  |  |
| Age <65 (n=3,869) | Ref | 0.80 (0.42, 1.53) | 0.73 (0.42, 1.27) | 1.29 (0.78, 2.14) | 0.06 | 1.22 (1.02, 1.46) | 0.73 |
| Age 65+ (n=4,169) | Ref | 1.28 (0.98, 1.67) | 1.43 (1.11, 1.84) | 1.50 (1.13, 1.98) |  | 1.18 (1.06, 1.31) |  |
| *Adjusted for continuous age* |  |  |  |  |  |  |  |
| Age <65 (N=3,869) | Ref | 0.71 (0.37, 1.36) | 0.67 (0.40, 1.14) | 1.01 (0.61, 1.69) | 0.07 | 1.09 (0.91, 1.31) | 0.73 |
| Age 65+ (N=4,169) | Ref | 1.21 (0.91, 1.61) | 1.25 (0.98, 1.60) | 1.16 (0.89, 1.51) |  | 1.06 (0.96, 1.17) |  |
| *Unadjusted for continuous age* |  |  |  |  |  |  |  |
| Age <65 (n=1,314) | Ref | 0.53 (0.15, 1.84) | 1.08 (0.43, 2.68) | 1.35 (0.55, 3.32) | 0.59 | 1.11 (0.84, 1.46) | 0.88 |
| Age 65+ (n=1,519) | Ref | 0.96 (0.60, 1.52) | 1.13 (0.70, 1.83) | 1.20 (0.71, 2.04) |  | 1.13 (0.97, 1.33) |  |
| *Adjusted for continuous age* |  |  |  |  |  |  |  |
| Age <65 (N=1,314) | Ref | 0.51 (0.15, 1.73) | 1.07 (0.43, 2.66) | 1.12 (0.46, 2.78) | 0.69 | 1.06 (0.80, 1.40) | 0.91 |
| Age 65+ (N=1,519) | Ref | 0.84 (0.52, 1.34) | 0.96 (0.61, 1.50) | 0.99 (0.58, 1.67) |  | 1.07 (0.92, 1.26) |  |

All models were adjusted for categorical age, sex, race/ethnicity, education, income-poverty ratio, smoking status, pack-year, serum cotinine, alcohol consumption, and body mass index. Urinary cadmium models were also adjusted for urinary creatinine. Multiplicative interaction terms between cadmium and categorical age were also included in the models.

No effect modification by sex was observed in NHANES III nor in continuous NHANES (**Tables S3, S4**). Among non-smoking participants, similar associations between cadmium concentrations and AD/all-cause dementia were observed as in the overall samples (**Tables S5, S6**). In NHANES III, we observed that a doubling of urinary cadmium was associated with 5% lower cumulative incidence of all-cause dementia (SHR 0.95 [95% CI: 0.91, 0.99]), when considering mortality due to other causes as a competing risk (**Table S7**). Finally, the use of competing risk regressions in continuous NHANES did not change our results (**Table S8**).

## Discussion

We evaluated the associations between cadmium exposure and incidences of all-cause dementia and AD using two independent and nationally representative samples, NHANES III (n=6,122) and continuous NHANES (n= 8,038 for blood cadmium; n=2,833 for urinary cadmium), with up to 30 years of follow-up via Medicare claims linkage. The current study does not support our hypothesis, as we did not observe associations between cadmium exposures and all-cause dementia or AD incidence, and these findings were consistent when stratifying by sex, and subset to non-smokers.

Other longitudinal studies have failed to find significant associations between cadmium exposures and related outcomes. For example, Peng, et al 2017 evaluated the association between cadmium exposures and AD mortality risk in two independent NHANES samples. ^27^ In this study, an association was found between blood and urinary cadmium and AD mortality risk in NHANES 1999-2006 after a follow-up of up to 12.7 years. However, in NHANES 1988-1994, which assessed cadmium exposure solely through urinary measurements, no statistically significant association was observed for a follow-up of up to 23 years. Methodological issues regarding creatinine adjustment, and length of follow-up could have explained the discrepancy between the samples. The association between urinary cadmium and AD mortality risk was statistically significant in NHANES 1988-1994 when models were not adjusted for creatinine, and the follow up time was restricted to 12.7 years (comparable to NHANES 1999-2006).^27^ The only other known study on cadmium and dementia incidence risk found a positive association between urinary cadmium and all-cause dementia risk over a follow up time of 16.7 years.^22^ Our null findings could be partially explained by the relatively longer mean follow up times of up to 30-years for NHANES III (1988-1994) and up to 19 years for continuous NHANES (1999-2016), which may have introduced survival bias. Thus, under the assumption that cadmium exposures play an important role in dementia and AD onset, it is possible that people highly vulnerable to cadmium toxicity died before AD or dementia onset earlier in the follow up, leaving individuals in our cohorts less susceptible to its effects.

Although we did not observe associations between cadmium and AD or dementia in a population study, compelling biological and mechanistic data still exist. Cadmium has been linked to adverse effects on the central nervous system and to the neuropathology of AD. The neurotoxic mechanisms of cadmium include mitochondrial dysfunction, glycogen metabolism dysfunction, and neurotransmission signaling alterations, all characteristics observed in AD.^13^ These neurotoxic mechanisms promote oxidative stress, leading to neuroinflammation, neuronal apoptosis, and a higher risk for neurodegenerative diseases.^13, 14^ Cadmium also plays a role in the development of specific AD neuropathologies, such as the formation of plaques and neurofibrillary tangles. Cadmium induces the formation of senile plaques by promoting the production of amyloid-beta (Aβ) through the stimulation of amyloid-beta precursor protein (APP), and by downregulating enzymes involved in reducing Aβ levels in the brain, such as α-secretase and neutral endopeptidase.^41, 42^ Cadmium promotes the conformation and accumulation of total and hyperphosphorylated tau in neurofibrillary tangles^43, 44^ by blocking muscarinic M1 receptors, which are known to downregulate glycogen synthase kinase 3 beta (GSK-3β), and Acetylcholinesterase variant S (AChE-S).^43, 45^ The formation of Aβ proteins and tau filament formation, alongside with the overexpression of GSK-3β and Acetylcholinesterase variant S (AChE-S) have been related to apoptosis of basal forebrain cholinergic neurons, critical in learning and memory processes.^43, 45^

Recent evidence has emerged linking cadmium exposure and subclinical AD characteristics in humans. For example, the Beaver Dam Offspring Study observed an association between blood cadmium with increased blood neurofilament light, a nonspecific marker of neurodegeneration, over a 10-year period. This study also found cross-sectional associations between blood cadmium, and higher serum total tau concentrations in adult women and a lower serum mean Aβ42/Aβ40 in both sexes.^46^ This study suggests that cadmium might play a role on AD and dementia progression.

Given that there is a plausible biological link between cadmium exposure and dementia risk, several methodological factors should be considered to explain our null findings. First, our results may have been subject to outcome misclassification. Medicare claims datasets are rich sources of health outcomes information for epidemiologic research,^47^ and its linkage to other studies such as NHANES is a cost-effective approach to evaluate longitudinal associations in the US population.^48^ However, for health outcomes with long latency periods, Medicare clinical claims may not capture early pathological changes occurring years before clinical onset. Medicare dementia claims have an estimated sensitivity of 85% and specificity of 89%, whereas AD claims have a sensitivity of 64% and specificity of 95%.^48^ Thus, the use of Medicare-based clinical outcomes in our study may have led to outcome misclassification, potentially biasing our results towards the null.

Second, in our study there was a time difference between exposure assessment and risk follow-up at Medicare eligibility (>65 years of age or presence of other conditions before 65) for both NHANES III and continuous NHANES. Since for most individuals, Medicare eligibility starts at 65 years of age, this follow-up gap was larger for younger NHANES participants, potentially leading to differential exposure measurement error if the exposure measurement did not reflect the exposure levels when they entered the risk set at Medicare eligibility. This measurement error could have led to the underestimation of exposure, as younger NHANES participants were likely to have lower cadmium levels at the time of exposure assessment than when they entered the risk set,^49^ while exposure measurements obtained from participants aged ≥65 before they entered the risk set could reflect more accurate exposure levels. This eligibility discrepancy led us to perform an <65 and ≥65 years age-stratified analysis. In the population <65 years of age of NHANES III, urinary cadmium was associated with increased AD, and all-cause dementia incidence risk. Similarly, in individuals ≥ 65 years from continuous NHANES, blood cadmium was associated with both higher risk for AD and all-cause dementia incidence. However, these associations were no longer statistically significant when adjusting for continuous age, also an important risk factor for AD and dementia,^4^ suggesting residual confounding by age in the initial age-stratified models. Nevertheless, the potential for exposure measurement error remains due to age differences at exposure assessment, as well as to the use of a single urinary or blood cadmium measurements, as they may not accurately reflect long-term cadmium exposures.

Third, another limitation of the use of Medicare data linkage for outcome ascertainment is the time difference between biospecimen collection, interview, and medical examination in NHANES, and risk follow-up based on Medicare data (starting in 1999). The timing of AD and all-cause dementia incidence were not observed if these conditions occurred before Medicare eligibility. Thus, it is possible that health characteristics at Medicare enrollment differ from those measured in NHANES, resulting in residual confounding. Additionally, this left truncation could have resulted in the underestimation of all-cause and AD dementia risk in our study. This contrasts with what was observed in the Multi-Ethnic Study of Atherosclerosis study which collected outcome information consistently from 2000-2002 through 2018 and found a positive association between urinary cadmium and all-cause dementia risk.^22^

Fourth, in NHANES III (1988-1994), the time gap between exposure assessment and outcome ascertainment was least five years and up to 11 years, potentially resulting in a higher proportion of individuals who developed all-cause dementia or AD before 1999, and were therefore censored, compared to continuous NHANES. This may have biased associations between cadmium and dementia incidence in NHANES III towards the null. Similarly, our analysis in continuous NHANES did not include around 30% of Medicare beneficiaries^47^ enrolled in a Medicare Part C Advantage plan. These, and other study design differences between NHANES III and continuous NHANES could explain the contrasting results we observed by sample in the age-stratified models. Finally, the presence of competing risks could lead to biased associations, where individuals die before having the chance to develop dementia. However, in our Fine-Grey analysis we observed that the consideration of competing risks had no effect on AD or all-cause dementia incidence.

Despite the limitations discussed above, our study has several strengths such as the use of two independent samples, each with a large sample size representative of the US, including a wide range of ages and multiple racial and ethnicity groups. Moreover, Medicare data linkage allowed for longitudinal analyses of the association between cadmium and AD/dementia incidence across a follow up of up to 20 years. We also considered two exposure biomarkers simultaneously, blood cadmium, which has an estimated half-life of 75 to 128 days,^50^ and urinary cadmium, which reflects long-term exposure with a half-life spanning from a few years given to decades. Considering diverse biomarkers of cadmium exposure allows for a more detailed assessment of cadmium exposure.

In two nationally representative samples, Medicare-linked NHANES III and continuous NHANES, we did not observe associations between cadmium exposure and dementia incidence. Although this study did not support our hypothesis, it provided evidence on the complexity of studying long latency diseases such as dementia and AD. Our results underline the need for further studies to elucidate the role of environmental exposures in dementia incidence.

## Acknowledgments

We thank Dieudonne Nahigombeye at the Centers for Disease Control and Prevention for his assistance.

## Author contributions

Erika Walker: data curation, software, formal analysis, visualization, writing- original draft, writing- review and editing

Yanelli Rodríguez-Carmona: visualization, writing- original draft, writing- review and editing

Xin Wang: data curation, software, writing- review and editing

Bhramar Mukherjee: writing- review and editing

Laura Arboleda-Merino: validation, writing- review and editing

Wei Hao: writing- review and editing

Hiroko Dodge: writing- review and editing

Roger L. Albin: writing- review and editing

Henry L. Paulson: writing- review and editing

Sung Kyun Park: conceptualization, methodology, supervision, project administration, funding acquisition, writing- review and editing

Kelly M. Bakulski: conceptualization, methodology, supervision, project administration, funding acquisition, writing- review and editing

All authors have read and approved the final manuscript.

## Statements and declarations

## NCHS disclaimer

The findings and conclusions in this paper are those of the author(s) and do not necessarily represent the views of the National Center for Health Statistics Research Data Center or the Centers for Disease Control and Prevention.

## Ethical considerations

Data collection for NHANES was approved by the NCHS Research Ethics Review Board. Analysis of de-identified data from the survey is exempt from the federal regulations for the protection of human research participants. Analysis of restricted data through the NCHS Research Data Center is also approved by the NCHS ERB. The current analysis of the existing Medicare-linked NHANES data has been approved by the University of Michigan Institutional Review Board (HUM00194918).

## Consent to participate

NHANES participants provided informed consent to the NCHS.

## Data availability

NHANES datasets are publicly available online through the NCHS. Access to linked Medicare datasets is restricted and requires an application process.

## References

1. Alzheimer’s Association. 2025 Alzheimer’s disease facts and figures. Alzheimer’s & Dementia 2025; 21: e70235. DOI: 10.1002/alz.70235.

2. Nichols E, et al. Estimation of the global prevalence of dementia in 2019 and forecasted prevalence in 2050: an analysis for the Global Burden of Disease Study 2019. Lancet Public Health 2022; 7: e105–e125. 2022/01/10. DOI: 10.1016/S2468-2667(21)00249-8.

3. Krishnamurthy HK, Jayaraman V, Krishna K, et al. An overview of the genes and biomarkers in Alzheimer’s disease. Ageing Res Rev 2025; 104: 102599. 2024/11/30. DOI: 10.1016/j.arr.2024.102599.

4. Eid A, Mhatre I and Richardson JR. Gene-environment interactions in Alzheimer’s disease: A potential path to precision medicine. Pharmacol Ther 2019; 199: 173–187. 20190312. DOI: 10.1016/j.pharmthera.2019.03.005.

5. Livingston G, Huntley J, Liu KY, et al. Dementia prevention, intervention, and care: 2024 report of the Lancet standing Commission. Lancet 2024; 404: 572–628. 2024/08/04. DOI: 10.1016/S0140-6736(24)01296-0.

6. Althobaiti NA. Heavy metals exposure and Alzheimer’s disease: Underlying mechanisms and advancing therapeutic approaches. Behav Brain Res 2025; 476: 115212. 2024/08/27. DOI: 10.1016/j.bbr.2024.115212.

7. Schaefer HR, Dennis S and Fitzpatrick S. Cadmium: Mitigation strategies to reduce dietary exposure. J Food Sci 2020; 85: 260–267. 2020/01/21. DOI: 10.1111/1750-3841.14997.

8. Zheng W. Toxicology of choroid plexus: special reference to metal-induced neurotoxicities. Microsc Res Tech 2001; 52: 89–103. 2001/01/03. DOI: 10.1002/1097-0029(20010101)52:1<89::AID-JEMT11>3.0.CO;2-2.

9. Takeda A, Takefuta S, Ijiro H, et al. 109Cd transport in rat brain. Brain Res Bull 1999; 49: 453–457. 1999/09/14. DOI: 10.1016/s0361-9230(99)00080-5.

10. Branca JJV, Maresca M, Morucci G, et al. Effects of Cadmium on ZO-1 Tight Junction Integrity of the Blood Brain Barrier. Int J Mol Sci 2019; 20 2019/12/05. DOI: 10.3390/ijms20236010.

11. Tjalve H, Henriksson J, Tallkvist J, et al. Uptake of manganese and cadmium from the nasal mucosa into the central nervous system via olfactory pathways in rats. Pharmacol Toxicol 1996; 79: 347–356. 1996/12/01. DOI: 10.1111/j.1600-0773.1996.tb00021.x.

12. Bondier JR, Michel G, Propper A, et al. Harmful effects of cadmium on olfactory system in mice. Inhal Toxicol 2008; 20: 1169–1177. 2008/10/28. DOI: 10.1080/08958370802207292.

13. Arruebarrena MA, Hawe CT, Lee YM, et al. Mechanisms of Cadmium Neurotoxicity. Int J Mol Sci 2023; 24 2023/12/09. DOI: 10.3390/ijms242316558.

14. Wen S and Wang L. Cadmium neurotoxicity and therapeutic strategies. J Biochem Mol Toxicol 2024; 38: e23670. 2024/03/04. DOI: 10.1002/jbt.23670.

15. Bakulski KM, Seo YA, Hickman RC, et al. Heavy Metals Exposure and Alzheimer’s Disease and Related Dementias. J Alzheimers Dis 2020; 76: 1215–1242. 2020/07/12. DOI: 10.3233/JAD-200282.

16. Basun H, Lind B, Nordberg M, et al. Cadmium in blood in Alzheimer’s disease and non-demented subjects: results from a population-based study. Biometals 1994; 7: 130–134. 1994/04/01. DOI: 10.1007/BF00140482.

17. Hart RP, Rose CS and Hamer RM. Neuropsychological effects of occupational exposure to cadmium. J Clin Exp Neuropsychol 1989; 11: 933–943. 1989/12/01. DOI: 10.1080/01688638908400946.

18. Nordberg M, Winblad B and Basun H. Cadmium concentration in blood in an elderly urban population. Biometals 2000; 13: 311–317. 2001/03/15. DOI: 10.1023/a:1009268123320.

19. Ciesielski T, Bellinger DC, Schwartz J, et al. Associations between cadmium exposure and neurocognitive test scores in a cross-sectional study of US adults. Environ Health 2013; 12: 13. 2013/02/06. DOI: 10.1186/1476-069X-12-13.

20. Gao S, Jin Y, Unverzagt FW, et al. Trace element levels and cognitive function in rural elderly Chinese. J Gerontol A Biol Sci Med Sci 2008; 63: 635–641. 2008/06/19. DOI: 10.1093/gerona/63.6.635.

21. Li H, Wang Z, Fu Z, et al. Associations between blood cadmium levels and cognitive function in a cross-sectional study of US adults aged 60 years or older. BMJ Open 2018; 8: e020533. 2018/04/15. DOI: 10.1136/bmjopen-2017-020533.

22. Domingo-Relloso A, McGraw KE, Heckbert SR, et al. Urinary Metal Levels, Cognitive Test Performance, and Dementia in the Multi-Ethnic Study of Atherosclerosis. JAMA Netw Open 2024; 7: e2448286. 20241202. DOI: 10.1001/jamanetworkopen.2024.48286.

23. Liu H, Su L, Chen X, et al. Higher blood cadmium level is associated with greater cognitive decline in rural Chinese adults aged 65 or older. Sci Total Environ 2021; 756: 144072. 20201126. DOI: 10.1016/j.scitotenv.2020.144072.

24. Yang Y, Yang W, Cao W, et al. Association of Toxic Metal Exposure with Cognitive Function in Elderly Chinese: Potential Modification by Difficulties Falling Asleep. Biol Trace Elem Res 2025 2025/08/13. DOI: 10.1007/s12011-025-04752-5.

25. Wei Y, Zhou YF, Xiao L, et al. Associations of Heavy Metals with Cognitive Function: An Epigenome-Wide View of DNA Methylation and Mediation Analysis. Ann Neurol 2024; 96: 87–98. 20240425. DOI: 10.1002/ana.26942.

26. Min JY and Min KB. Blood cadmium levels and Alzheimer’s disease mortality risk in older US adults. Environ Health 2016; 15: 69. 2016/06/16. DOI: 10.1186/s12940-016-0155-7.

27. Peng Q, Bakulski KM, Nan B, et al. Cadmium and Alzheimer’s disease mortality in U.S. adults: Updated evidence with a urinary biomarker and extended follow-up time. Environ Res 2017; 157: 44–51. 2017/05/17. DOI: 10.1016/j.envres.2017.05.011.

28. Chen S, Shen R, Shen J, et al. Association of blood cadmium with all-cause and cause-specific mortality in patients with hypertension. Front Public Health 2023; 11: 1106732. 20230704. DOI: 10.3389/fpubh.2023.1106732.

29. Jack CR, Jr., Andrews JS, Beach TG, et al. Revised criteria for diagnosis and staging of Alzheimer’s disease: Alzheimer’s Association Workgroup. Alzheimers Dement 2024; 20: 5143–5169. 2024/06/27. DOI: 10.1002/alz.13859.

30. Centers for Disease Control and Prevention. NHANES Questionnaires, Datasets, and Related Documentation, https://wwwn.cdc.gov/nchs/nhanes/default.aspx (2025, accessed August 15 2025).

31. Centers for Disease Control and Prevention. National Center for Health Statistics. NCHS Data Linkage. Restricted-Use NCHS-CMS Medicare, https://www.cdc.gov/nchs/data-linkage/medicare-restricted.htm (2023).

32. National Center for Health Statistics. Linkage Methods and Analytic Support for NCHS-CMS Medicare Data, https://www.cdc.gov/nchs/data-linkage/medicare-methods.htm (2024).

33. National Center for Health Statistics. NCHS Data Linked to NDI Mortality Files, https://www.cdc.gov/nchs/data-linkage/mortality.htm (2024).

34. Vacchi-Suzzi C, Kruse D, Harrington J, et al. Is Urinary Cadmium a Biomarker of Long-term Exposure in Humans? A Review. Curr Environ Health Rep 2016; 3: 450–458. DOI: 10.1007/s40572-016-0107-y.

35. Centers for Disease Control and Prevention. National Center for Health Statistics. NHANES III Laboratory data file documentation. 2006.

36. Centers for Disease Control and Prevention. National Center for Health Statistics. Laboratory Data - Continuous NHANES. 2025.

37. Centers for Disease Control and Prevention. National Center for Health Statistics. National Health and Nutrition Examination Survey 1999-2000 Data Documentation, Codebook, and Frequencies. Metals - Urine (LAB06HM), https://wwwn.cdc.gov/Nchs/Data/Nhanes/Public/1999/DataFiles/LAB06HM.htm (2023, accessed January 13 2025).

38. Centers for Disease Control and Prevention. National Center for Health Statistics. National Health and Nutrition Examination Survey 2001-2002 Data Documentation, Codebook, and Frequencies. Metals - Urine (L06HM_B), https://wwwn.cdc.gov/Nchs/Data/Nhanes/Public/2001/DataFiles/L06HM_B.htm#URDUCD (2023, accessed January 13 2025).

39. Centers for Medicare & Medicaid Services. Chronic Conditions Data Warehouse, https://www2.ccwdata.org/condition-categories-chronic.

40. Fine JP and Gray RJ. A Proportional Hazards Model for the Subdistribution of a Competing Risk. Journal of the American Statistical Association 1999; 94: 496–509. DOI: 10.1080/01621459.1999.10474144.

41. Li X, Lv Y, Yu S, et al. The effect of cadmium on Abeta levels in APP/PS1 transgenic mice. Exp Ther Med 2012; 4: 125–130. 2012/10/13. DOI: 10.3892/etm.2012.562.

42. Notarachille G, Arnesano F, Calo V, et al. Heavy metals toxicity: effect of cadmium ions on amyloid beta protein 1-42. Possible implications for Alzheimer’s disease. Biometals 2014; 27: 371–388. 2014/02/22. DOI: 10.1007/s10534-014-9719-6.

43. Del Pino J, Zeballos G, Anadon MJ, et al. Cadmium-induced cell death of basal forebrain cholinergic neurons mediated by muscarinic M1 receptor blockade, increase in GSK-3beta enzyme, beta-amyloid and tau protein levels. Arch Toxicol 2016; 90: 1081–1092. 2015/06/01. DOI: 10.1007/s00204-015-1540-7.

44. Jiang LF, Yao TM, Zhu ZL, et al. Impacts of Cd(II) on the conformation and self-aggregation of Alzheimer’s tau fragment corresponding to the third repeat of microtubule-binding domain. Biochim Biophys Acta 2007; 1774: 1414–1421. 2007/10/09. DOI: 10.1016/j.bbapap.2007.08.014.

45. Del Pino J, Zeballos G, Anadon MJ, et al. Muscarinic M1 receptor partially modulates higher sensitivity to cadmium-induced cell death in primary basal forebrain cholinergic neurons: A cholinesterase variants dependent mechanism. Toxicology 2016; 361-362: 1–11. 2016/07/06. DOI: 10.1016/j.tox.2016.06.019.

46. Schubert CR, Paulsen AJ, Pinto AA, et al. Effect of Neurotoxin Exposure on Blood Biomarkers of Neurodegeneration and Alzheimer Disease. Alzheimer Dis Assoc Disord 2023; 37: 310–314. 20230913. DOI: 10.1097/WAD.0000000000000579.

47. Mues KE, Liede A, Liu J, et al. Use of the Medicare database in epidemiologic and health services research: a valuable source of real-world evidence on the older and disabled populations in the US. Clin Epidemiol 2017; 9: 267–277. 2017/05/24. DOI: 10.2147/CLEP.S105613.

48. Taylor DH, Jr., Ostbye T, Langa KM, et al. The accuracy of Medicare claims as an epidemiological tool: the case of dementia revisited. J Alzheimers Dis 2009; 17: 807–815. 2009/06/23. DOI: 10.3233/JAD-2009-1099.

49. Olsson IM, Bensryd I, Lundh T, et al. Cadmium in blood and urine--impact of sex, age, dietary intake, iron status, and former smoking--association of renal effects. Environ Health Perspect 2002; 110: 1185–1190. DOI: 10.1289/ehp.021101185.

50. Jarup L, Rogenfelt A, Elinder CG, et al. Biological half-time of cadmium in the blood of workers after cessation of exposure. Scand J Work Environ Health 1983; 9: 327–331. 1983/08/01. DOI: 10.5271/sjweh.2404.

