## Supplemental Material for "Cadmium Exposure and Incidence of All-Cause Dementia and Alzheimer’s Disease in US Adults"

Supplemental Materials

**Table S1.** International Classification of Disease (ICD) codes used to define Alzheimer’s disease (AD) and all-cause dementia in the Medicare Chronic Conditions Summary File. Claims with these ICD codes from inpatient services, skilled nursing facilities, home health agencies, hospital outpatient services, or carriers were defined as cases.

|  | **ICD-9** | **ICD-10** |
| --- | --- | --- |
| **AD** | **331.0:** Alzheimer’s disease | **G30.0:** Alzheimer’s disease with early onset  **G30.1:** Alzheimer’s disease with late onset  **G30.8:** Other Alzheimer’s disease  **G30.9:** Alzheimer’s disease, unspecified |
| **All-cause dementia** | **331.0:** Alzheimer’s disease  **331.11:** Pick’s disease (frontotemporal dementia)  **331.19:** Other frontotemporal dementia  **331.2:** Senile degeneration of brain  **331.7:** Cerebral degeneration in diseases classified elsewhere  **290.0:** Senile dementia, uncomplicated  **290.10:** Presenile dementia, uncomplicated  **290.11:** Presenile dementia with delirium  **290.12:** Presenile dementia with delusional features  **290.13:** Presenile dementia with depressive features  **290.20:** Senile dementia with delusional features  **290.21:** Senile dementia with depressive features  **290.3:** Senile dementia with delirium  **290.40:** Vascular dementia, uncomplicated  **290.41:** Vascular dementia with delirium  **290.42:** Vascular dementia with delusions  **290.43:** Vascular dementia with depressed mood  **294.0:** Amnestic disorder in conditions classified elsewhere  **294.10:** Dementia in conditions classified elsewhere without behavioral disturbance  **294.11:** Dementia in conditions classified elsewhere with behavioral disturbance  **294.20:** Dementia, unspecified, without behavioral disturbance  **294.21:** Dementia unspecified, with behavioral disturbance  **294.8:** Other persistent mental disorders due to conditions classified elsewhere  **797:** Senility without mention of psychosis | **F01.50:** Vascular dementia, unspecified severity, without behavioral disturbance, psychotic disturbance, mood disturbance, and anxiety  **F01.51:** Vascular dementia with behavioral disturbance  **F02.80:** Dementia in other diseases classified elsewhere, unspecified severity, without behavioral disturbance, psychotic disturbance, mood disturbance, and anxiety  **F02.81:** Dementia in other diseases classified elsewhere with behavioral disturbance  **F03.90:** Unspecified dementia, unspecified severity, without behavioral disturbance, psychotic disturbance, mood disturbance, and anxiety  **F03.91:** Unspecified dementia with behavioral disturbance  **F04:** Amnestic disorder due to known physiological condition  **F05:** Delirium due to known physiological condition  **F06.1:** Catatonic disorder due to known physiological condition  **F06.8:** Other specified mental disorders due to known physiological condition  **G13.8:** Systemic atrophy primarily affecting central nervous system in other diseases classified elsewhere  **G30.0:** Alzheimer’s disease with early onset  **G30.1:** Alzheimer’s disease with late onset  **G30.8:** Other Alzheimer’s disease  **G30.9:** Alzheimer’s disease, unspecified **G31.01:** Pick’s disease (frontotemporal dementia)  **G31.09:** Other frontotemporal neurocognitive disorder  **G31.1:** Senile degeneration of brain, not elsewhere classified  **G31.2:** Degeneration of nervous system due to alcohol  **G94:** Other disorders of brain in diseases classified elsewhere  **R41.81:** Age-related cognitive decline  **R54:** Age-related physical debility |

**Table S2.** Characteristics of included versus excluded participants from NHANES III with Medicare claims linkage (N=8,792) and from continuous NHANES participants (N=13,866)

|  | **NHANES III** | | | |  | **Continuous NHANES** | | | |
| --- | --- | --- | --- | --- | --- | --- | --- | --- | --- |
|  | **All**  N=8,792 | **Included**  N=6,122 | **Excluded**  N =2,670 | ***p*-value** |  | **All**  N=13,866 | **Included**  N=8,038 | **Excluded**  N=5,828 | ***p* -value** |
| **Continuous variables, mean (SE)** | | | | |  |  |  |  |  |
| **Urinary cadmium, ug/L** | 0.77 (0.02) | 0.76 (0.02) | 0.81 (0.03) | 0.13 |  | 0.50 (0.02) | 0.49 (0.03) | 0.54 (0.02) | 0.11 |
| Missing | 1,116 | 0 | 1,116 |  |  | 9,674 | 5,207 | 4,467 |  |
| **Blood cadmium, ug/L** | - | - | - | - |  | 0.56 (0.01) | 0.55 (0.01) | 0.60 (0.01) | **0.0004** |
| Missing |  |  |  |  |  | 2,211 | 0 | 2,211 |  |
| **Follow-up time, years** | 19.98 (0.28) | 20.35 (0.26) | 18.89 (0.43) | **<0.0001** |  | 8.63 (0.11) | 9.46 (0.12) | 7.24 (0.16) | **<0.0001** |
| **Age, years** | 54.66 (0.45) | 53.86 (0.46) | 56.96 (0.57) | **<0.0001** |  | 65.29 (0.16) | 64.13 (0.19) | 67.23 (0.20) | **<0.0001** |
| **Pack-years** | 14.60 (0.43) | 14.29 (0.46) | 15.62 (0.85) | 0.15 |  | 15.50 (0.32) | 14.03 (0.37) | 18.12 (0.54) | **<0.0001** |
| Missing | 359 | 0 | 359 |  |  | 352 | 0 | 352 |  |
| **Serum cotinine, ng/mL** | 70.09 (2.50) | 70.47 (2.65) | 67.51 (5.50) | 0.61 |  | 47.14 (1.50) | 45.38 (1.78) | 50.92 (2.38) | 0.05 |
| Missing | 1,389 | 0 | 1,389 |  |  | 1,418 | 0 | 1,418 |  |
| **Body mass index, kg/m^2^** | 27.32 (0.13) | 27.36 (0.14) | 27.17 (0.21) | 0.43 |  | 28.89 (0.09) | 29.10 (0.13) | 28.78 (0.09) | **0.02** |
| Missing | 822 | 0 | 822 |  |  | 1,043 | 0 | 1,043 |  |
| **Urinary creatinine, mg/dL** | 113.24 (1.50) | 113.78 (1.64) | 110.18 (2.38) | 0.18 |  | 106.84 (0.85) | 106.24 (0.98) | 107.99 (1.40) | 0.28 |
| Missing | 1,147 | 0 | 1,147 |  |  | 1,018 | 67 | 951 |  |
| **Categorical variables, n (%)** | | | | |  |  |  |  |  |
| **Sex** |  |  |  | **<0.0001** |  |  |  |  | **0.01** |
| Male | 3,975 (45.47%) | 2,797 (46.88%) | 1,178 (41.44%) |  |  | 6,911 (46.30%) | 3,822 (45.39%) | 3,089 (47.83%) |  |
| Female | 4,817 (54.53%) | 3,325 (53.13%) | 1,492 (58.56%) |  |  | 6,955 (53.70%) | 4,216 (54.61%) | 2,739 (52.17%) |  |
| **Race/ethnicity** |  |  |  | **<0.0001** |  |  |  |  | **<0.0001** |
| Non-Hispanic White | 4,394 (81.37%) | 3,130 (82.54%) | 1,264(78.01%) |  |  | 7,543 (79.59%) | 4,618(81.71%) | 2,925(76.05%) |  |
| Non-Hispanic Black | 2,077 (8.93%) | 1,398 (8.16%) | 679 (11.15%) |  |  | 2,648 (8.55%) | 1,403 (7.51%) | 1,245 (10.30%) |  |
| Mexican American | 1,982 (3.52%) | 1,352 (3.37%) | 630 (3.94%) |  |  | 2,054 (3.70%) | 1,168 (3.40%) | 886 (4.20%) |  |
| Other Hispanic | - | - | - | - |  | 887 (3.44%) | 488 (3.35%) | 399 (3.59%) |  |
| Other | 339 (6.18%) | 242 (5.93%) | 97 (6.90%) |  |  | 734 (4.72%) | 361 (4.04%) | 373 (5.85%) |  |
| **Education** |  |  |  | **<0.0001** |  |  |  |  | **<0.0001** |
| Less than high school | 3,823 (26.82%) | 2,491 (24.54%) | 1,332 (33.60%) |  |  | 4,566 (20.27%) | 2,300 (17.10%) | 2,266 (25.60%) |  |
| High school graduate | 3,696 (51.19%) | 2,743 (52.36%) | 953 (47.74%) |  |  | 6,551 (52.63%) | 3,945 (53.43%) | 2,606 (51.27%) |  |
| College and above | 1,195 (21.98%) | 888 (23.10%) | 307 (18.66%) |  |  | 2,723 (27.10%) | 1,793 (29.47%) | 930 (23.12%) |  |
| Missing | 78 | 0 | 78 |  |  | 26 | 0 | 26 |  |
| **Income-poverty ratio** |  |  |  | **0.002** |  |  |  |  | **<0.0001** |
| ≤ 1 | 1,452 (9.01%) | 1,077 (8.34%) | 375 (11.68%) |  |  | 2,150 (9.88%) | 1,191 (8.32%) | 959 (13.07%) |  |
| >1 | 6,458 (90.99%) | 5,045 (91.66%) | 1,413 (88.32%) |  |  | 10,586 (90.12%) | 6,847 (91.68%) | 3,739 (86.93%) |  |
| Missing | 882 | 0 | 882 |  |  | 1,130 | 0 | 1,130 |  |
| **Smoking status** |  |  |  | **0.0003** |  |  |  |  | **<0.0001** |
| Never | 4,010 (42.44%) | 2,896 (43.86%) | 1,114 (38.38%) |  |  | 6,528 (46.88%) | 4,087 (50.13%) | 2,441 (41.44%) |  |
| Former | 2,793 (33.63%) | 1,923 (33.45%) | 870 (34.13%) |  |  | 5,203 (37.87%) | 2,739 (34.75%) | 2,464 (43.09%) |  |
| Current | 1,982 (23.93%) | 1,303 (22.69%) | 679 (27.50%) |  |  | 2,120 (15.25%) | 1,212 (15.12%) | 908 (15.47%) |  |
| Missing | 7 | 0 | 7 |  |  | 15 | 0 | 15 |  |
| **Alcohol consumption** |  |  |  | 0.86 |  |  |  |  | 0.09 |
| 0 drink/day | 6,639 (86.98%) | 5,413 (87.03%) | 1,226 (86.68%) |  |  | 9,033 (86.21%) | 7,023 (85.78%) | 2,010 (87.88%) |  |
| ≥1 drink/day | 859 (13.02%) | 709 (12.97%) | 150 (13.32%) |  |  | 1,246 (13.79%) | 1,015 (14.22%) | 231 (12.12%) |  |
| Missing | 1,294 | 0 | 1,294 |  |  | 3,587 | 0 | 3,587 |  |
| Note: Survey weighted means and standard errors were calculated for continuous variables. Unweighted frequencies and survey weighted percentages were calculated for categorical variables. Survey weighted t-tests were used to test continuous variables, and survey weighted Chi-square test was used for categorical variables. | | | | | | | | | |

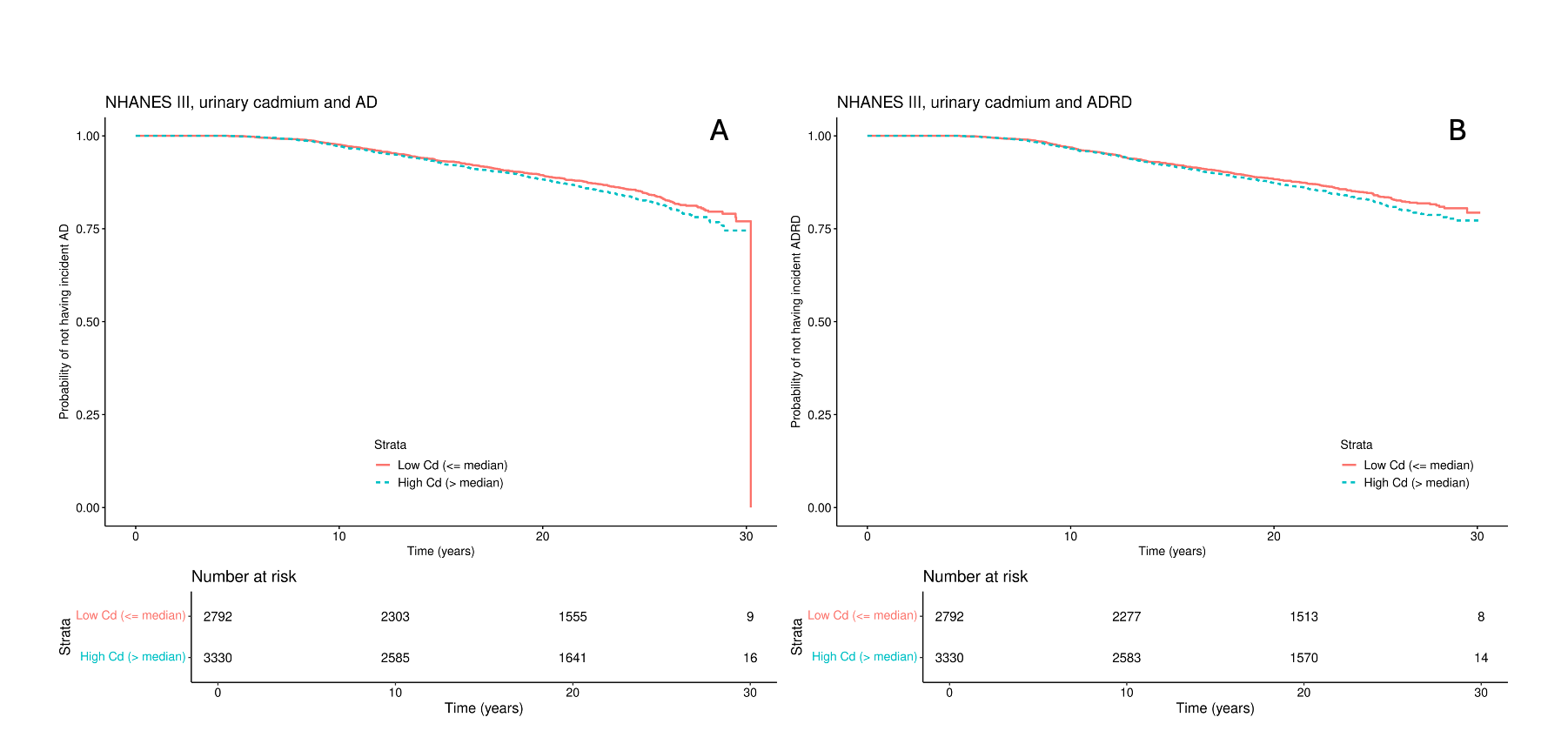

**Figure S1**. Unweighted Kaplan-Meier (K-M) curves showing the probability of remaining free from 1) Alzheimer’s disease (AD) or 2) All-cause dementia, stratified by median concentrations of urinary cadmium in NHANES III.

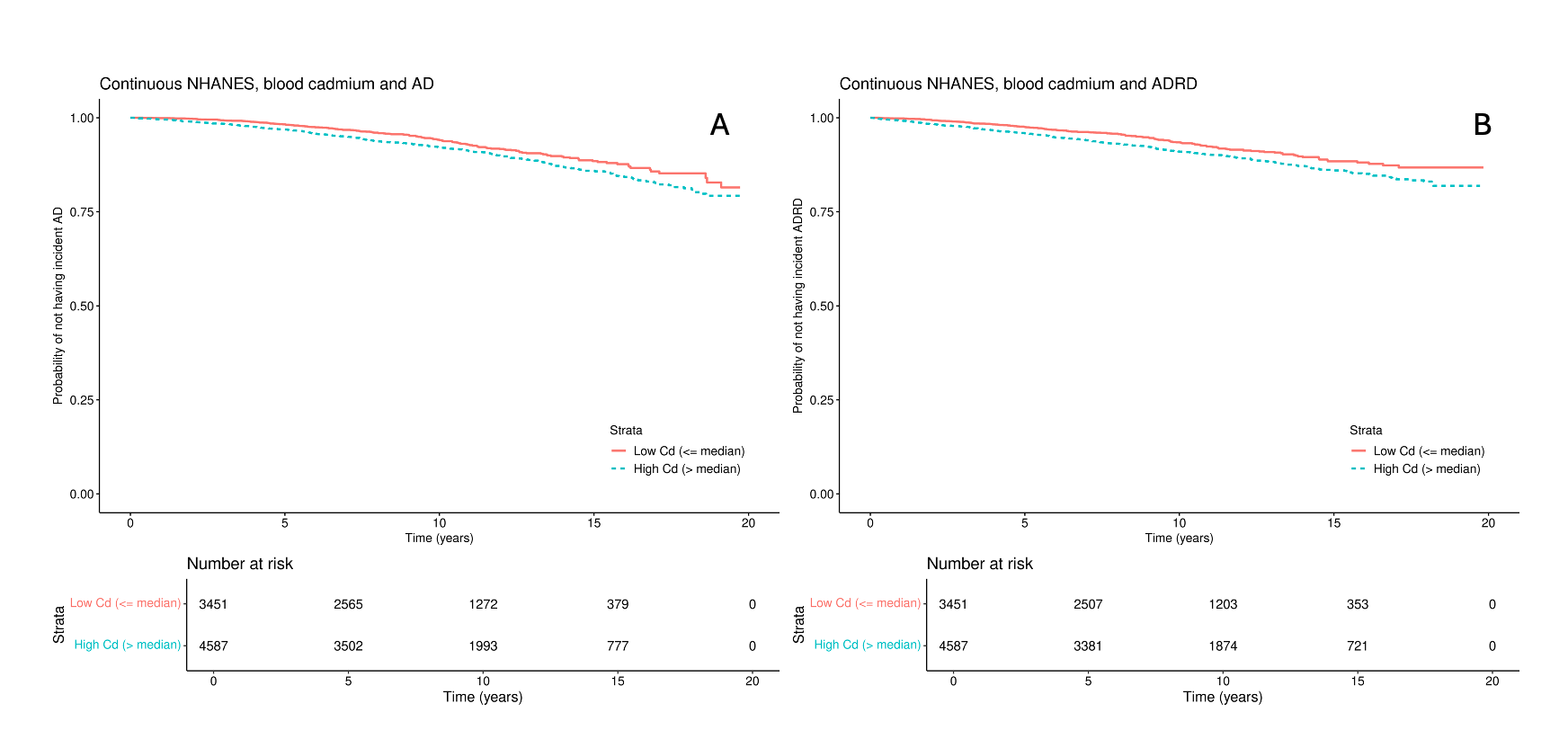

**Figure S2**. Unweighted Kaplan-Meier (K-M) curves showing the probability of remaining free from 1) Alzheimer’s disease (AD) or 2) All-cause dementia, stratified by median concentrations of blood cadmium in continuous NHANES.

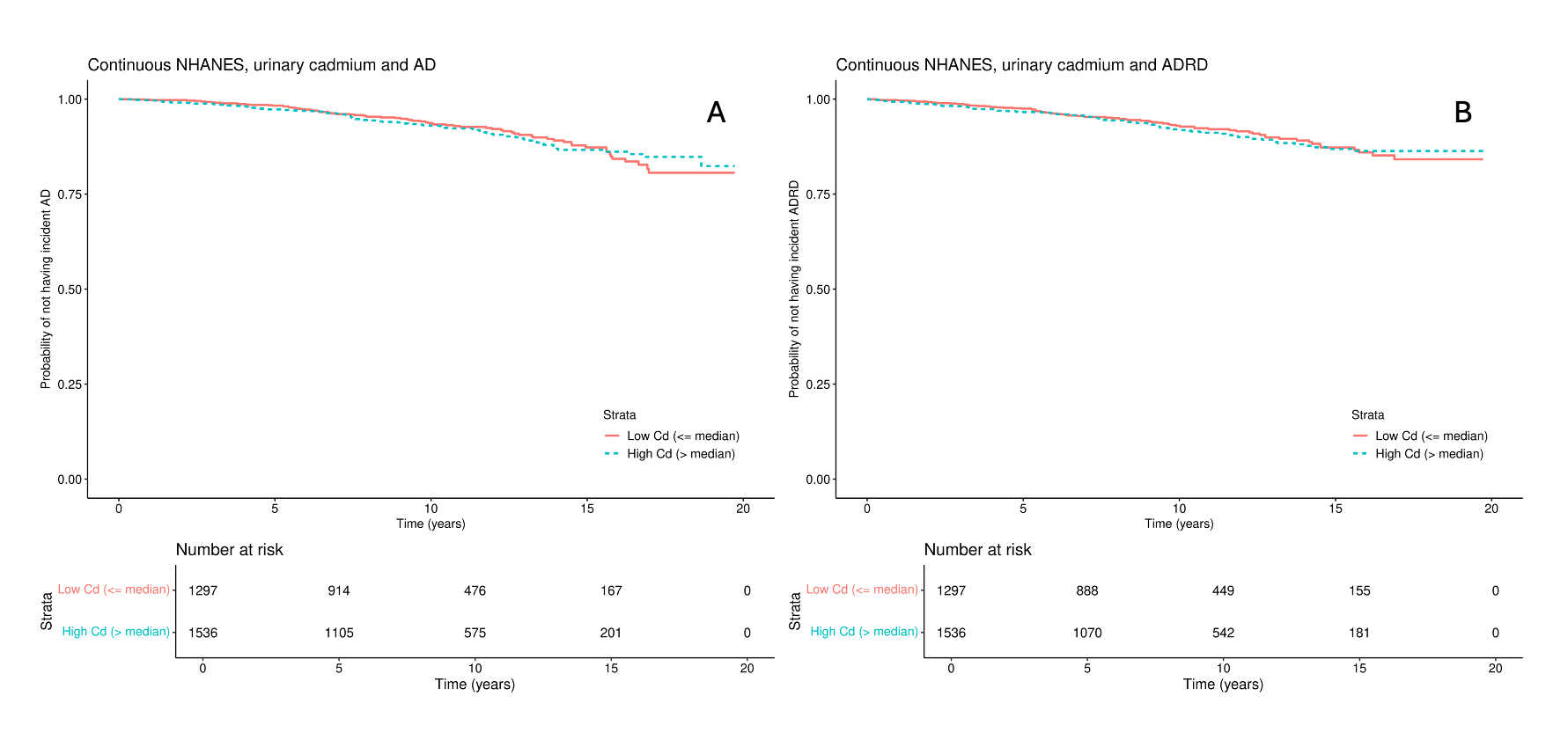

**Figure S3**. Unweighted Kaplan-Meier (K-M) curves showing the probability of remaining free from 1) Alzheimer’s disease (AD) or 2) All-cause dementia, stratified by median concentrations of urinary cadmium in continuous NHANES

**Table S3.** Effect modification of associations of cadmium exposure and incident Alzheimer’s disease (AD) and all-cause dementia by sex in NHANES III (N=6,122)

|  | **Quartiles of urinary cadmium** | | | | |  | **Continuous urinary cadmium** | |
| --- | --- | --- | --- | --- | --- | --- | --- | --- |
|  | **Q1** | **Q2**  HR (95% CI) | **Q3**  HR (95% CI) | **Q4**  HR (95% CI) | *p* for interaction |  | **Per doubling**  HR (95% CI) | *p* for interaction |
| **AD** | | | | | | | | |
| Women | Ref | 0.86 (0.56, 1.33) | 0.74 (0.48, 1.15) | 1.00 (0.58, 1.73) | 0.57 |  | 1.04 (0.93, 1.16) | 0.14 |
| Men | Ref | 1.09 (0.68, 1.74) | 0.83 (0.52, 1.31) | 0.83 (0.48, 1.45) |  |  | 0.95 (0.87, 1.04) |  |
| **All-cause dementia** | | | | | | | | |
| Women | Ref | 0.99 (0.74, 1.33) | 0.86 (0.65, 1.15) | 1.05 (0.74, 1.50) | 0.80 |  | 1.02 (0.95, 1.09) | 0.62 |
| Men | Ref | 0.98 (0.72, 1.33) | 0.94 (0.65, 1.35) | 0.91 (0.62, 1.35) |  |  | 1.00 (0.93, 1.07) |  |
| Notes: Quartiles of cadmium concentrations are based on weighted distributions.  All models were adjusted for age, sex, race/ethnicity, education, income-poverty ratio, smoking status, pack-year, serum cotinine, alcohol consumption, body mass index, and urinary creatinine. Multiplicative interaction terms between cadmium and sex were also included in the models. | | | | | | | | |

**Table S4.** Effect modification of associations of cadmium exposure and incident Alzheimer’s disease (AD) and all-cause dementia by sex in continuous NHANES.

|  | **Quartiles of blood or urinary cadmium concentrations** | | | | |  | **Continuous blood or urinary cadmium concentrations** | |
| --- | --- | --- | --- | --- | --- | --- | --- | --- |
|  | **Q1** | **Q2**  HR (95% CI) | **Q3**  HR (95% CI) | **Q4**  HR (95% CI) | *p* for interaction |  | **Per doubling**  HR (95% CI) | *p* for interaction |
| **AD** | | | | | | | | |
| Blood cadmium | | | | | | | | |
| Women | Ref | 1.25 (0.69, 2.25) | 1.33 (0.79, 2.25) | 1.16 (0.68, 1.98) | 0.95 |  | 1.09 (0.92, 1.30) | 0.49 |
| Men | Ref | 1.06 (0.63, 1.78) | 1.09 (0.69, 1.71) | 1.00 (0.60, 1.67) |  |  | 1.00 (0.82, 1.22) |  |
| Urinary cadmium | | | | | | | | |
| Women | Ref | 0.91 (0.44, 1.89) | 0.98 (0.44, 2.18) | 1.02 (0.46, 2.25) | 0.92 |  | 1.10 (0.88, 1.37) | 0.83 |
| Men | Ref | 0.84 (0.33, 2.09) | 0.88 (0.34, 2.30) | 0.70 (0.20, 2.48) |  |  | 1.06 (0.75, 1.49) |  |
| **All-cause dementia** | | | | | | | | |
| Blood cadmium | | | | | | | | |
| Women | Ref | 1.28 (0.81, 2.01) | 1.54 (1.07, 2.20) | 1.32 (0.91, 1.91) | **0.02** |  | 1.10 (0.98, 1.24) | 0.19 |
| Men | Ref | 0.99 (0.70, 1.40) | 0.76 (0.54, 1.06) | 0.96 (0.68, 1.37) |  |  | 0.99 (0.86, 1.14) |  |
| Urinary cadmium | | | | | | | | |
| Women | Ref | 0.80 (0.47, 1.39) | 1.04 (0.61, 1.77) | 0.96 (0.56, 1.66) | 0.88 |  | 1.06 (0.91, 1.22) | 0.65 |
| Men | Ref | 0.75 (0.39, 1.41) | 0.94 (0.50, 1.77) | 1.13 (0.55, 2.32) |  |  | 1.11 (0.88, 1.40) |  |
| Notes: Quartiles of cadmium concentrations are based on weighted distributions.  All models were adjusted for age, sex, race/ethnicity, education, income-poverty ratio, smoking status, pack-year, serum cotinine, alcohol consumption, and body mass index. Multiplicative interaction terms between lead and sex were also included in the models. | | | | | | | | |

**Table S5:** Associations between cadmium and incident Alzheimer’s disease (AD) and all-cause dementia among non-smokers in NHANES III (N=2,896). Quartiles (Q) of cadmium concentrations are based on weighted distributions.

|  | | **Quartiles of urinary cadmium** | | | |  | **Continuous urinary cadmium** | |  |
| --- | --- | --- | --- | --- | --- | --- | --- | --- | --- |
|  | **Q1** | | **Q2**  HR (95% CI) | **Q3**  HR (95% CI) | **Q4**  HR (95% CI) | ***p*-trend** |  | **Per doubling**  HR (95% CI) | ***p*** |
| **AD** | | | | | | | | | |
| Cases / Quartile | 102/825 | | 103/830 | 105/743 | 78/498 |  |  |  |  |
| Estimates | Ref | | 1.00 (0.64, 1.56) | 0.98 (0.61, 1.57) | 1.16 (0.67, 2.02) | 0.68 |  | 1.06 (0.95, 1.18) | 0.32 |
| **All-cause dementia** | | | | | | | | | |
| Cases / Quartile | 207/825 | | 217/830 | 206/743 | 143/498 |  |  |  |  |
| Estimates | Ref | | 1.07 (0.81, 1.43) | 1.10 (0.84, 1.44) | 1.03 (0.73, 1.46) | 0.74 |  | 1.03 (0.96, 1.10) | 0.39 |
| All models were adjusted for age, sex, race/ethnicity, education, income-poverty ratio, pack-year, serum cotinine, alcohol consumption, body mass index, and urinary creatinine. | | | | | | | | | |

**Table S6:** Associations between cadmium and incident AD and All-cause dementia among non-smokers in continuous NHANES (N=5,507).

|  | **Quartiles of blood and urinary cadmium** | | | | |  | **Continuous blood and urinary cadmium** | |
| --- | --- | --- | --- | --- | --- | --- | --- | --- |
|  | **Q1** | **Q2**  HR (95% CI) | **Q3**  HR (95% CI) | **Q4**  HR (95% CI) | ***p-*trend** |  | **Per doubling**  HR (95% CI) | ***p*** |
| **AD** | |  |  |  |  |  |  |  |
| Blood cadmium (n=4,087) | |  |  |  |  |  |  |  |
| Range ug/L | 0.07-0.26 | 0.27-0.39 | 0.40-0.59 | 0.60-8.40 |  |  |  |  |
| Cases / Quartile | 59/1,235 | 71/1,028 | 125/1,201 | 76/623 |  |  |  |  |
| Estimates | Ref | 1.11 (0.72, 1.72) | 1.26 (0.84, 1.88) | 1.22 (0.75, 1.97) | 0.34 |  | 1.09 (0.91, 1.31) | 0.35 |
| Urinary cadmium (n=1,420) | |  |  |  |  |  |  |  |
| Range ug/L | 0.01-0.16 | 0.16-0.31 | 0.32-0.57 | 0.57-37.78 |  |  |  |  |
| Cases / Quartile | 25/423 | 28/371 | 30/359 | 25/267 |  |  |  |  |
| Estimates | Ref | 0.80 (0.38, 1.68) | 0.93 (0.37, 2.35) | 1.21 (0.42, 3.53) | 0.75 |  | 1.11 (0.81, 1.50) | 0.52 |
| **All-cause dementia** | |  |  |  |  |  |  |  |
| Blood cadmium (n=4,087) | |  |  |  |  |  |  |  |
| Range ug/L | 0.07-0.26 | 0.27-0.39 | 0.40-0.59 | 0.60-8.40 |  |  |  |  |
| Cases / Quartile | 131/1,235 | 149/1,028 | 252/1,201 | 149/623 |  |  |  |  |
| Estimates | Ref | 1.08 (0.78, 1.49) | 1.26 (0.99, 1.61) | 1.14 (0.83, 1.57) | 0.21 |  | 1.08 (0.94, 1.24) | 0.29 |
| Urinary cadmium (n=1,420) | |  |  |  |  |  |  |  |
| Range ug/L | 0.01-0.16 | 0.16-0.31 | 0.32-0.57 | 0.57-37.78 |  |  |  |  |
| Cases / Quartile | 52/423 | 50/371 | 66/359 | 57/267 |  |  |  |  |
| Estimates | Ref | 0.64 (0.36, 1.13) | 0.88 (0.50, 1.56) | 1.01 (0.54, 1.90) | 0.82 |  | 1.03 (0.87, 1.22) | 0.74 |
| Notes: Quartiles of cadmium concentrations are based on weighted distributions.  All models were adjusted for age, sex, race/ethnicity, education, income-poverty ratio, pack-year, serum cotinine, alcohol consumption, and body mass index. Urinary cadmium models were adjusted further for urinary creatinine. | | | | | | | | |

**Table S7:** Associations between cadmium and incident Alzheimer’s disease (AD) and all-cause dementia in NHANES III using Fine-Gray competing risk regression model (N=6,122)

|  | **Quartiles of urinary cadmium** | | | | |  | **Continuous urinary cadmium** | |
| --- | --- | --- | --- | --- | --- | --- | --- | --- |
|  | **Q1** | **Q2**  SHR (95% CI) | **Q3**  SHR (95% CI) | **Q4**  SHR (95% CI) | ***p*-trend** |  | **Per doubling**  SHR (95% CI) | ***p*** |
| **AD^a^** | Ref | 0.93 (0.74, 1.17) | 0.95 (0.74, 1.21) | 0.95 (0.71, 1.27) | 0.82 |  | 0.97 (0.92, 1.03) | 0.37 |
| **All-cause dementia^b^** | Ref | 0.87 (0.73, 1.03) | 0.83 (0.70, 1.00) | 0.85 (0.68, 1.05) | 0.14 |  | 0.95 (0.91, 0.99) | **0.02** |
| Notes: Quartiles of cadmium concentrations are based on weighted distributions.  The survey design was not included in Fine-Gray competing risk regression models.  All models were adjusted for age, sex, race/ethnicity, education, income-poverty ratio, smoking status, pack-year, serum cotinine, alcohol consumption, body mass index, and urinary creatinine. The competing risk outcome was coded as incident AD or all-cause dementia, mortality (competing risk), or censored.  ^a^AD models: N=743 AD cases and 2,656 mortality cases.  ^b^All-cause dementia models: N=1,508 All-cause dementia cases and 2,014 mortality cases. | | | | | | | | |

**Table S8:** Associations between cadmium and incident Alzheimer’s disease (AD) and all-cause dementia in continuous NHANES using Fine-Gray competing risk regression model. Quartiles (Q) of cadmium concentrations are based on weighted distributions

|  | **Quartiles of cadmium** | | | | |  | **Continuous cadmium** | |
| --- | --- | --- | --- | --- | --- | --- | --- | --- |
|  | **Q1** | **Q2**  SHR (95% CI) | **Q3**  SHR (95% CI) | **Q4**  SHR (95% CI) | ***p*-trend** |  | **Per doubling**  SHR (95% CI) | ***p*** |
| **AD^a^** |  |  |  |  |  |  |  |  |
| Blood cadmium | Ref | 1.11 (0.83, 1.48) | 1.26 (0.97, 1.64) | 1.00 (0.74, 1.34) | 0.89 |  | 1.00 (0.89, 1.11) | 0.95 |
| Urinary cadmium | Ref | 0.94 (0.59, 1.50) | 0.99 (0.60, 1.63) | 1.00 (0.55, 1.80) | 0.95 |  | 1.06 (0.89, 1.25) | 0.54 |
| **All-cause dementia^b^** |  |  |  |  |  |  |  |  |
| Blood cadmium | Ref | 1.12 (0.92, 1.36) | 1.18 (0.99, 1.41) | 1.08 (0.88, 1.32) | 0.45 |  | 1.04 (0.96, 1.12) | 0.38 |
| Urinary cadmium | Ref | 0.85 (0.61, 1.19) | 1.03 (0.73, 1.46) | 1.01 (0.68, 1.50) | 0.66 |  | 1.04 (0.93, 1.17) | 0.46 |
| The survey design was not included in Fine-Gray competing risk regression models.  All models were adjusted for age, sex, race/ethnicity, education, income-poverty ratio, smoking status, pack-year, serum cotinine, alcohol consumption, and body mass index. The competing risk outcome was coded as incident AD or all-cause dementia, mortality (competing risk), or censored.  ^a^AD models: N=587 AD cases and 2,135 mortality cases.  ^b^All-cause dementia models: N=1,260 All-cause dementia cases and 1,683 mortality cases. | | | | | | | | |
